# Exome analysis of 22,319 individuals links extremely rare CNVs and 22q11.21 dosage to Alzheimer’s risk

**DOI:** 10.1101/2024.10.28.24314051

**Authors:** Olivier Quenez, Catherine Schramm, Kévin Cassinari, Aude Nicolas, Joan Groeneveld, Guillaume Huguet, Benjamin Grenier-Boley, Marc Hulsman, G. Bragi Walters, Itziar de Rojas, Anne Rovelet-Lecrux, Sébastien Feuillette, Laetitia Miguel, Anne-Claire Richard, Stéphane Rousseau, Shahzad Ahmad, Najaf Amin, Philippe Amouyel, Olivia Belbin, Céline Bellenguez, Claudine Berr, Paola Bossù, Femke Bouwman, Jose Bras, Jordi Clarimon, Antonio Daniele, Jean-François Dartigues, Stéphanie Debette, Jean-François Deleuze, Nicola Denning, Oriol Dols-Icardo, Cornelia M. van Duijn, Juan Fortea, Nick C. Fox, Ruth Frikke-Schmidt, Daniela Galimberti, Roberta Ghidoni, Vilmantas Giedraitis, Johan J. P. Gille, Detelina Grozeva, Rita Guerreiro, Edna Grünblatt, John Hardy, Steffi G. Riedel-Heller, Mikko Hiltunen, Clive Holmes, Jakub Hort, Holger Hummerich, M. Arfan Ikram, M. Kamran Ikram, Martin Ingelsson, Iris E. Jansen, Amit Kawalia, Robert Kraaij, Patrick G. Kehoe, Marc Lathrop, Morgane Lacour, Afina W. Lemstra, Alberto Lleó, Lauren Luckcuck, Marcel M. A. M. Mannens, Rachel Marshall, Carlo Masullo, Simon Mead, Patrizia Mecocci, Alexandre de Mendonça, Alun Meggy, Shima Mehrabian, Merel O. Mol, Kevin Morgan, Alexandre Morin, Benedetta Nacmias, Penny J. Norsworthy, Robert Olaso, Florence Pasquier, Pau Pastor, Fabrizio Piras, Julius Popp, Alfredo Ramirez, Rachel Raybould, Richard Redon, Marcel J. T. Reinders, Fernando Rivadeneira, Jeroen G. J. van Rooij, Natalie S. Ryan, Salha Saad, Pascual Sanchez-Juan, Nikolaos Scarmeas, Philip Scheltens, Jonathan M. Schott, Davide Seripa, Daoud Sie, Rebecca Sims, Erik A. Sistermans, Sandro Sorbi, Kristel Sleegers, Resie van Spaendonk, John C. van Swieten, Niccolo’ Tesi, Betty M Tijms, Magda Tsolaki, André G. Uitterlinden, Jort Vijverberg, Pieter Jelle Visser, Michael Wagner, Julie Williams, Aline Zarea, EADB Consortium, Emmanuelle Génin, Henne Holstege, Daniel F. Gudbjartsson, David Wallon, Magalie Lecourtois, Maria Victoria Fernandez, Hreinn Stefansson, Sébastien Jacquemont, Jean-Charles Lambert, Sven J. van der Lee, Camille Charbonnier, Gaël Nicolas

## Abstract

Copy number variants (CNVs), defined as deletions or duplications of genomic segments >100 bp, are major contributors to human disease, yet their role in non-monogenic Alzheimer disease (AD) remains poorly characterized. We analyzed rare CNVs from 22,319 exomes, including 4,150 early-onset AD (EOAD), 8,519 late-onset AD and 9,650 unaffected controls. EOAD cases showed increased burdens of rare CNVs, particularly rare deletions within AD-related genes. Loss-of-function analyses implicated deletions in *ABCA1* and *ABCA7* and identified *CTSB* as a candidate *locus*. Exome-wide gene-level dosage analysis highlighted 18 genes across five *loci*, including chr22q11.21, where deletions were restricted to EOAD and duplications enriched in controls. Replication in 33,992 cases and 362,305 controls confirmed AD-risk reduction in 22q11.21 duplication carriers (mega-analysis dosage OR(SCARF2)=0.34, p=5.52×10^-7^). *SCARF2* overexpression increased amyloid-β uptake *in vitro*, supporting a functional link. These findings highlight CNVs as contributors to AD risk and identify 22q11.21 dosage as a strong genetic determinant.

## INTRODUCTION

The etiology of Alzheimer disease (AD) includes autosomal dominant AD (ADAD), caused by pathogenic variants in *PSEN1, PSEN2* or *APP* and representing <1% of all cases, and non-monogenic (or complex AD), combining genetic and non-genetic factors. Genetic factors play a prominent role in non-monogenic AD, and the etiology of non-monogenic early-onset AD (EOAD, onset ≤65 years) shows an even greater genetic component^1^.

To date, most identified AD genetic determinants are single nucleotide variants (SNVs) and short insertions/deletions (indels), from ADAD-causing pathogenic variants to a large diversity of risk factors in non-monogenic AD. Among the latter ones, common variants all show modest odds ratios (ORs) below 1.5^2^, except the *APOE-ε4* allele with ORs ranging from ∼3 to ∼14 depending on ancestry and zygosity, despite a high frequency (9-23%)^1^. Multiple rare (allele frequency<1%) coding variants in five genes (*SORL1, TREM2, ABCA7, ABCA1* and *ATP8B4*), mostly identified by exome sequencing, are associated with higher odds ratios (ORs), ranging from 1.5 to over 20 and thus contribute significantly to the risk of AD in carriers^1, 3^. Recent findings from exome or genome sequencing studies show that (i) the rarer the variants, the higher the ORs, and (ii) rare variants with strong effects on AD risk are significantly enriched in EOAD cases compared to late-onset (LOAD) patients^3, 4^. In contrast with AD risk-increasing variants, only a handful of variants have been significantly associated with strong effects on AD risk reduction (OR<0.7), so far.

Copy number variants (CNVs, deletions or duplications of >100-bp genomic DNA regions) represent a variant type with a major role in multiple diseases^5-7^. In AD, CNVs have been less studied than SNVs and indels. *APP* locus duplications on chromosome 21 are typical examples of CNVs causing ADAD^8, 9^, accounting for ∼9% of all ADAD cases (update of ref.^10^). Importantly, they also underscore why Trisomy-21 patients develop AD with full penetrance by age 70^11^. In addition, they demonstrate that increased expression of the Amyloid β (Aβ) precursor protein (APP) is sufficient to cause AD, further supporting the amyloid cascade stating that Aβ aggregation is a mandatory triggering factor of AD. Ultra-rare in-frame exon9/exon9-10 deletions of *PSEN1* also cause ADAD^12^, while a few common CNVs, detected by DNA chips, have emerged as candidate risk factors^13, 14^. However, none of them reached genome-wide significance. Regarding rare CNVs, a handful of studies using DNA chips were published, on a limited number of EOAD patients,^15, 16^ before the advent of next generation sequencing. DNA chips can only detect CNVs larger than 50-100 kb, thus missing smaller CNVs, which show a remarkable diversity of rare events^17^.

Bioinformatics advances now enable the detection of rare CNVs from sequencing data. Recently, structural variants – including CNVs – were detected from genome sequencing data of 6,525 AD cases (average age at onset: 74.6 years, mainly LOAD patients) and 6,855 controls from the Alzheimer Disease Sequencing Project (ADSP) consortium^18, 19^. A moderate burden of (coding and non-coding) ultra-rare CNVs was associated with AD status, overall. These structural variants included a few examples of individual CNVs with a potential impact in the respective carriers, including one partial deletion of *SORL1* and five partial deletions of *ABCA7*, each identified in one patient. However, no gene-level association was assessed and no CNV was significantly associated with AD, likely due to limited recurrence of CNVs and also possibly because this cohort was not enriched in EOAD cases, limiting power. This power limitation is intrinsic to rare variant association studies, and this is even more pronounced for CNVs: whereas each individual carries hundreds of rare coding SNVs or indels, only one rare coding CNV is typically observed per genome, on average^17^. This makes it necessary to analyze larger datasets and/or enrich them in patients with higher suspected genetic component, as EOAD patients. Despite the development of genome sequencing datasets, a large proportion of the data available so far is made of exomes. While multiple algorithms are often combined for genome sequencing, exome-based approaches relying on specific read-depth tools have been shown to achieve high accuracy for rare coding CNVs without requiring multi-caller strategies^20, 21^. For example, we previously assessed the performances of a CNV-calling workflow based on the CANOES tool^22^, a clinical-grade validated CNV workflow that showed good sensitivity (87.25 %) and positive predictive value (86.4%-90.2%) for rare CNVs from exome sequencing data^23^. This makes large exome datasets particularly well suited for gene-level CNV association analyses

To identify CNVs associated with AD risk, we used one of the largest sequencing datasets available worldwide, i.e. the combination of the European and American consortia into the ADES-ADSP dataset (Alzheimer Disease European Sequencing – Alzheimer Disease Sequencing Project) as it is enriched in EOAD patients (32.7%), thus enhancing power for detecting CNVs with a moderate-to-strong effect. Importantly, this dataset contains a large proportion of exome sequencing data (68.6% after QC), allowing the use of a single caller with a high level of accuracy. We performed an AD case–control study using CNV calling from 22,319 exomes and focused on rare CNVs. We first searched for pathogenic CNV carriers, then performed burden analyses at the protein-coding genome level and after restriction to AD gene lists, and finally performed a genome-wide case-control association study at the gene level, different from any previous study in the field. Prioritized loci were further assessed by performing an integrated loss-of-function (LOF) analysis combining short truncating SNV/indels with CNV-deletions, a multi-ethnic replication analysis, and *in vitro* cellular assays.

## RESULTS

We included all 22,319 exomes from the combined ADES-ADSP dataset as described in ref.^3^, made of European and American cohorts, and proceeded to harmonized CNV detection, sample and variant quality control and filtration, followed by variant interpretation in a Mendelian setting and then, case-control association analyses (see online methods and detailed methods in supplementary information.

### Detection of CNVs from exome sequencing data of 22,319 individuals

Out of 22,319 exomes initially included, 22,110 passed QC (4093 EOAD cases, 8458 LOAD cases (late-onset AD) and, 9559 unaffected controls) (Table 1, Figure 1, supplementary information, Tables S1-S3), totaling 261,190 CNVs. The first step consisted in searching for pathogenic CNVs in monogenic dementia genes. We identified 17 carriers of such a pathogenic CNV, all but one with EOAD. Nine cases were novel, as 8 were previously reported^10, 12, 24-26^. Overall, we detected 10 *APP* duplications (mean age at onset (AAO) = 49.6 years, min-max: [43-58]), one in-frame *PSEN1* deletion (AAO=56 years), and six *MAPT* duplications, leading to a primary tauopathy differential diagnosis (mean AAO = 58.2, min-max: [45-87]) (Table 2). Carriers of pathogenic variants were excluded from further case-control analyses.

**Table 1.**
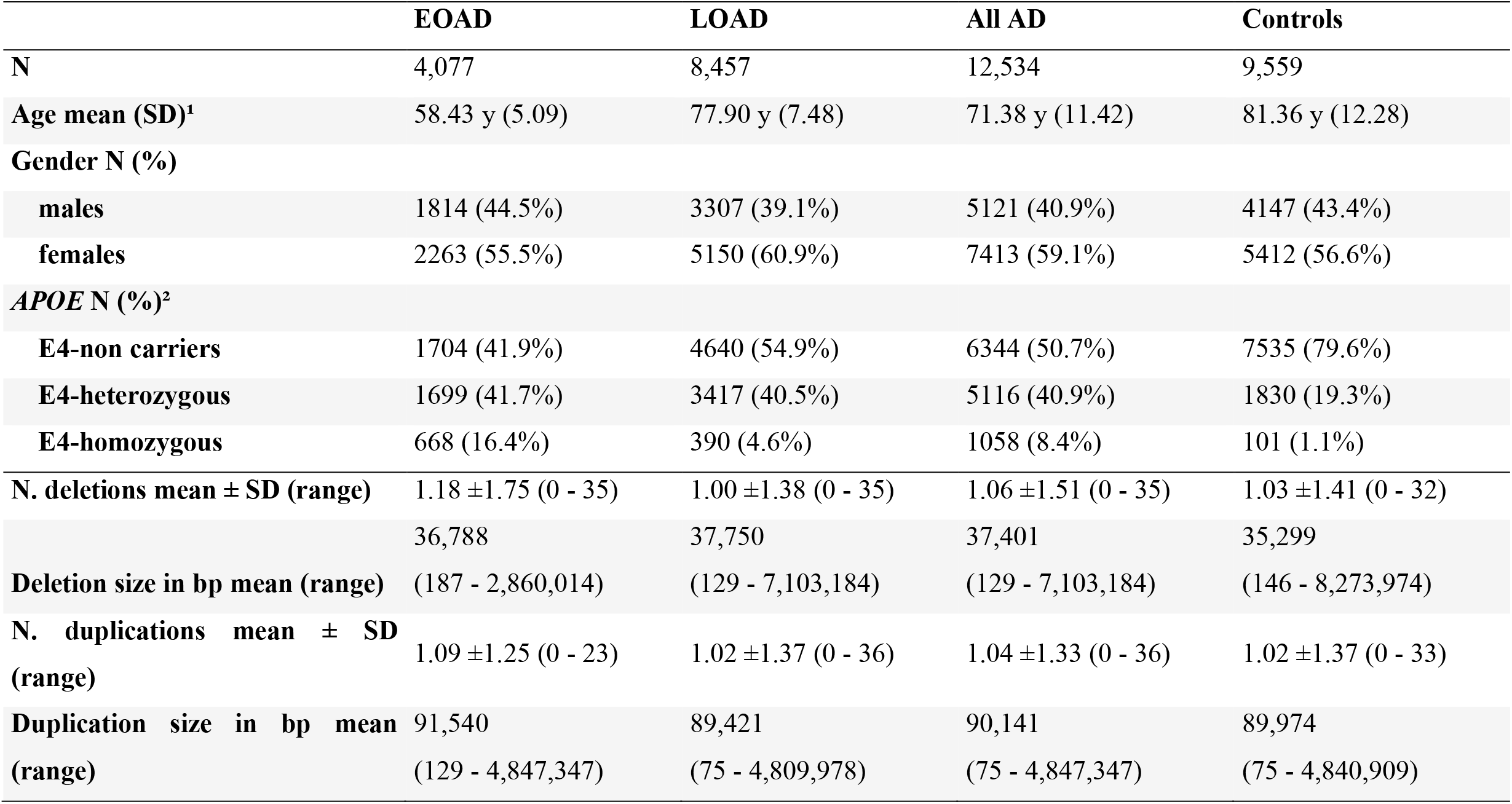
Description of individuals included in case-control analyses. N: number, y: years, SD: standard deviation, bp: base pairs, EOAD: early onset Alzheimer disease, LOAD: late onset Alzheimer disease ^1^AAO for cases and age at last exam/inclusion for controls. Age is missing for 373 LOAD and 256 controls ^2^APOE genotype is missing for 6 EOAD, 10 LOAD and 93 controls

|  | **EOAD** | **LOAD** | **All AD** | **Controls** |
| --- | --- | --- | --- | --- |
| **N** | 4,077 | 8,457 | 12,534 | 9,559 |
| **Age mean (SD)<sup>1</sup>** | 58.43 y (5.09) | 77.90 y (7.48) | 71.38 y (11.42) | 81.36 y (12.28) |
| **Gender N (%)** |  |  |  |  |
| **males** | 1814 (44.5%) | 3307 (39.1%) | 5121 (40.9%) | 4147 (43.4%) |
| **females** | 2263 (55.5%) | 5150 (60.9%) | 7413 (59.1%) | 5412 (56.6%) |
| **APOE N (%)<sup>2</sup>** |  |  |  |  |
| **E4-non carriers** | 1704 (41.9%) | 4640 (54.9%) | 6344 (50.7%) | 7535 (79.6%) |
| **E4-heterozygous** | 1699 (41.7%) | 3417 (40.5%) | 5116 (40.9%) | 1830 (19.3%) |
| **E4-homozygous** | 668 (16.4%) | 390 (4.6%) | 1058 (8.4%) | 101 (1.1%) |
| **N. deletions mean ± SD (range)** | 1.18 ±1.75 (0 - 35) | 1.00 ±1.38 (0 - 35) | 1.06 ±1.51 (0 - 35) | 1.03 ±1.41 (0 - 32) |
|  | 36,788 | 37,750 | 37,401 | 35,299 |
| **Deletion size in bp mean (range)** | (187 - 2,860,014) | (129 - 7,103,184) | (129 - 7,103,184) | (146 - 8,273,974) |
| **N. duplications mean ± SD (range)** | 1.09 ±1.25 (0 - 23) | 1.02 ±1.37 (0 - 36) | 1.04 ±1.33 (0 - 36) | 1.02 ±1.37 (0 - 33) |
|  | 91,540 | 89,421 | 90,141 | 89,974 |
| **Duplication size in bp mean (range)** | (129 - 4,847,347) | (75 - 4,809,978) | (75 - 4,847,347) | (75 - 4,840,909) |

**Table 2.** Mendelian pathogenic CNVs. AAO: Age at onset (in years), CNV: copy number variant, HGVS: Human Genome Variation Society; in bold: novel ^1^ reported in Le Guennec et al., Mol Psychiatry, 2017 ^2^ reported in Le Guennec et al., Neurobiol Dis, 2017 ^3^ reported in Lanoiselée et al., PLoS Med, 2017 ^4^ reported in Rovelet-Lecrux et al., Mol Psychiatry, 2015

| Sample | Cohort | AAO | Gender | CNV coordinate | HGVS nomenclature | Gene | Event |
| --- | --- | --- | --- | --- | --- | --- | --- |
| ALZ-0596-001<sup>1</sup> | ADES-FR | 49-50 | Male | chr17:43,679,169-44,145,099 | NC_000017.10:g.43585921_44159772dup | MAPT | complete duplication |
| ALZ-0441-005<sup>1</sup> | ADES-FR | 41-45 | Male | chr17:43,717,946-44,364,336 | NC_000017.10:g.43679455_44363695dup | MAPT | complete duplication |
| EXT-1114-001<sup>1</sup> | ADES-FR | 51-55 | Female | chr17:43,679,164-44,364,341 | NC_000017.10:g.43585926_44408484dup | MAPT | complete duplication |
| ROU-1373-001<sup>1</sup> | ADES-FR | 51-55 | Male | chr17:43,717,946-44,159,945 | NC_000017.10:g.43679455_44172015dup | MAPT | complete duplication |
| **SC45A00007a** | **PERADES** | **56-60** | **Female** | **chr17:43,679,204-44,159,910** | **NC_000017.10:g.43611648_44171922dup** | **MAPT** | **complete duplication** |
| **C-FHS-51259** | **ADSP** | **86-90** | **Female** | **chr17:43,898,720-44,101,538** | **NC_000017.10:g.43893949_44108840dup** | **MAPT** | **complete duplication** |
| EXT-0814-001<sup>3</sup> | ADES-FR | 51-55 | Female | chr21:22,370,838-28,338,704 | NC_000021.8:g.20058403_30248689dup | APP | complete duplication |
| EXT-0773-001<sup>3 4</sup> | ADES-FR | 41-45 | Male | chr21:26,946,242-31,066,399 | NC_000021.8:g.22910384_31233900dup | APP | complete duplication |
| EXT-0857-001<sup>3</sup> | ADES-FR | 56-60 | Male | chr21:26,946,246-28,338,704 | NC_000021.8:g.22910379_30248689dup | APP | complete duplication |
| **FR16640974** | **Netherlands Brain Bank** | **46-50** | **Male** | **chr21:22,370,838-28,338,827** | **NC_000021.8:g.20230358_30248689dup** | **APP** | **complete duplication** |
| **TE17A00020a_DS1** | **PERADES** | **41-45** | **Male** | **chr21:27,056,123-28,338,719** | **NC_000021.8:g.27012218_30248703dup** | **APP** | **complete duplication** |
| **20732** | **PERADES** | **51-55** | **Female** | **chr21:27,062,175-27,484,514** | **NC_000021.8:g.27056277_27512460dup** | **APP** | **complete duplication** |
| **CH32A00027a** | **PERADES** | **51-55** | **Female** | **chr21:27,066,065-28,338,719** | **NC_000021.8:g.27062287-30248703dup** | **APP** | **complete duplication** |
| **09D04764** | **ADC-Amsterdam** | **56-50** | **Male** | **chr21:26,541,838-29,645,932** | **NC_000021.8:g.25725527_30248655dup** | **APP** | **complete duplication** |
| **10D05060** | **ADC-Amsterdam** | **41-45** | **Female** | **chr21:27,253,167-27,542,996** | **NC_000021.8:g.27142399_27840733dup** | **APP** | **complete duplication** |
| **SID14617** | **AgeCoDe-UKBonn** | **46-50** | **Female** | **chr21:26,946,221-27,542,992** | **NC_000021.8:g.25725527_27840779dup** | **APP** | **complete duplication** |
| EXT-0313-001<sup>2</sup> | ADES-FR | 56-60 | Male | chr14:73,672,981-73,678,665 | NC_000014.8:g.73664857_73683810del | PSEN1 | deletion exon 9-10 |

**Figure 1:**
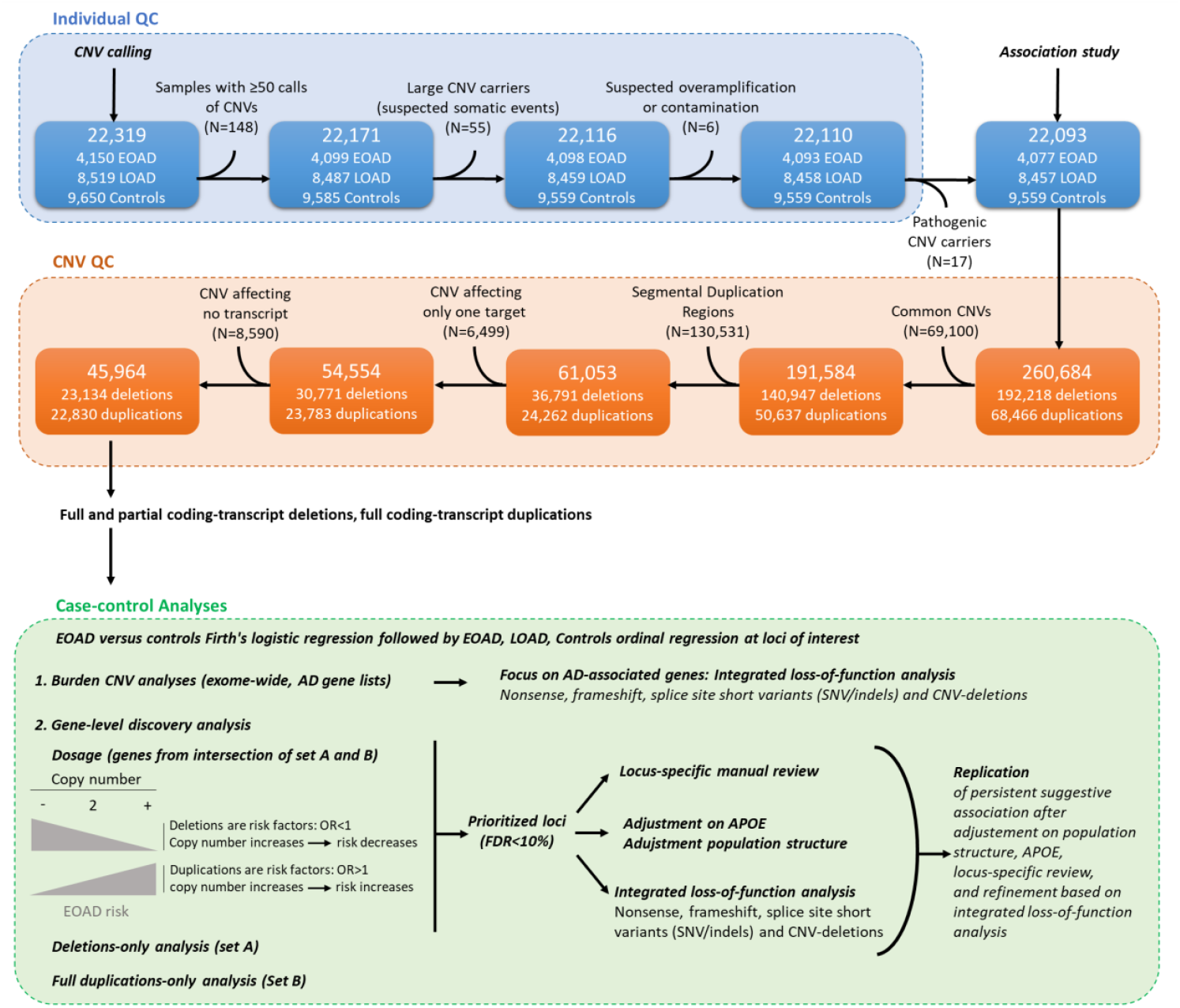
Summary of the study. This study included exome sequencing samples from the ADES-ADSP described in Holstege et al. 2022, restricted to individuals passing their quality control and being included in a CNV-calling-batch of 50+ individuals. QC: Quality Control; CNV: Copy Number Variation; EOAD: Early Onset Alzheimer Disease; LOAD: Late Onset Alzheimer Disease; OR: Odd-ratio Set A: non frequently deleted genes Set B: non frequently duplicated genes

### Burden CNV analyses in the protein-coding genome and focus on AD-related genes

We assessed if EOAD patients exhibit a higher burden of rare CNVs overall, as compared to controls, at two levels (Table S4). First, we performed an exome-wide burden CNV analysis across all protein-coding genes and showed that EOAD patients have a higher likelihood of carrying at least one deleted (set A, non-frequently deleted genes, see methods) or one duplicated (set B, non-frequently duplicated genes) gene than controls (deletions burden: OR=1.15 [1.06; 1.23], p=3×10^-4^; complete duplications burden: OR=1.10 [1.02; 1.19],p=0.0125). Second, restricting to AD-GWAS genes^2^, the difference was amplified for deletions (OR=2.67 [1.51; 4.74], p=8×10^-4^) and reversed for duplications (OR=0.58 [0.30; 1.04], p-value=0.0696) suggesting that some deletions in GWAS genes increase EOAD risk while some complete duplications could be protective (Table S4), and at-risk duplications in other genes may remain to be identified. Analysis of the Aβ-network list of genes^27^ indicated enrichment in deletions among EOAD cases (OR=1.44 [1.09; 1.90], p=0.0114, Table S4).

Then, to further assess the deletions-related signals, we performed a gene-based integrated loss-of-function (LOF) analysis on a list of known AD-associated genes, gathering short truncating variants (nonsense, frameshift, canonical splice site SNVs/indels) with CNV-deletions (complete and partial) (see online methods and supplementary information). We identified a significant enrichment of *ABCA1* (OR_EOAD_=5.77 [2.25 ; 17.06], p=2×10^-4^) and *ABCA7* (OR_EOAD_=2.29 [1.44 ; 3.65], p=6×10^-6^) LOF alleles with AD, now adding CNV-deletions to the known *ABCA7* association^28^, while the *CTSB* gene, mapping to a GWAS locus, emerged as a novel candidate (OR_EOAD_=5.03 [1.50 ; 20.71], p=8.9×10^-3^) (Tables S5, S6, additional results in supplementary information).

### Gene-level case-control discovery analysis

Then, we performed a gene-level rare CNV association study on the 22,093 samples remaining after exclusion of pathogenic CNV carriers. By Firth’s logistic regression, we compared the gene dosage (partial/complete deletions = decreased dosage, complete duplications = increased dosage) between EOAD cases and controls. In dosage analysis, ORs should be interpreted for an increase of gene copy by one, i.e. an OR >1 (resp. OR <1) means that duplications (resp. deletions) are associated with a higher disease risk (Figure 1, Figure 2, Table 3). Loci with False Discovery Rate (FDR) below 10% were then assessed in the whole cohort using an ordinal regression including EOAD, LOAD and controls (Table S7). Additional analyses included deletion-only (Table S8) or full duplication-only (Table S9) analyses, *APOE4*-adjusted (Table S10) and population structure-adjusted (Table S11) analyses. All analyses are transcript based, and results are represented per gene when several transcripts show identical results in a given gene.

**Table 3.**
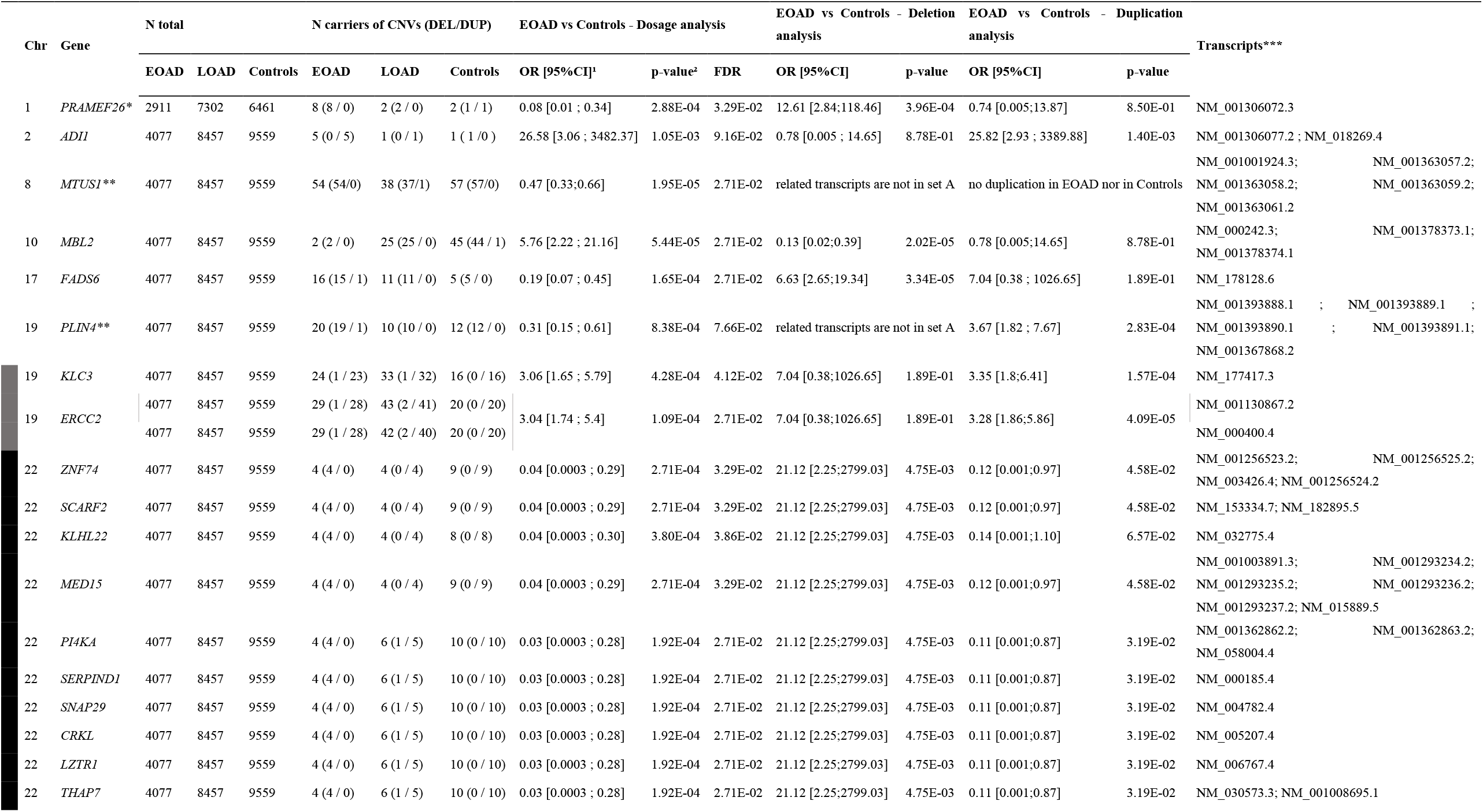
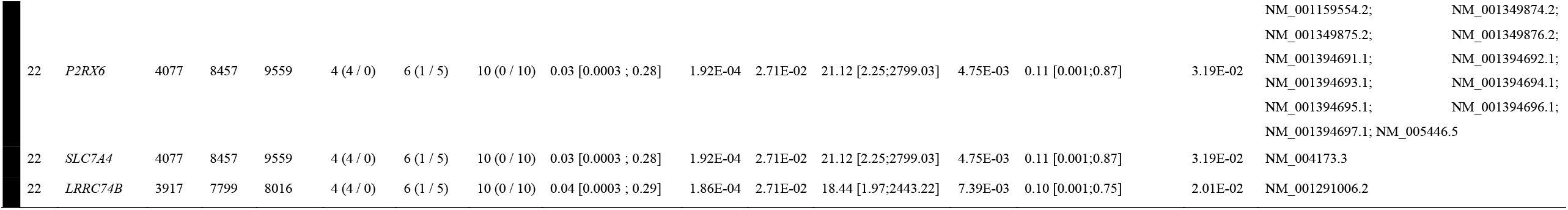
Results from EOAD versus Controls dosage analysis and subsequent analyses on deletions and duplications. Chr: chromosome; N: number. EOAD: early onset Alzheimer disease. LOAD: late onset Alzheimer disease ^1^OR should be interpreted for an increase of transcript copies by 1. i.e. a OR > 1 (resp. OR <1) means that duplications (resp. deletions) are associated with a higher disease risk ^2^non adjusted p-values. selected based on FRD < 10% ^*^gene overlapping with repeats or duplicated gene in the genome (despite CNVs not overlapping >50% with repeats) ^**^gene for which the signal in dosage analysis is driven by deletion whereas transcripts are not in set A ^***^ the table gathers transcript results on a single line per gene when several transcripts exhibit the same p-values Genes at the same locus are marked by a colored block on the left

**Figure 2.**
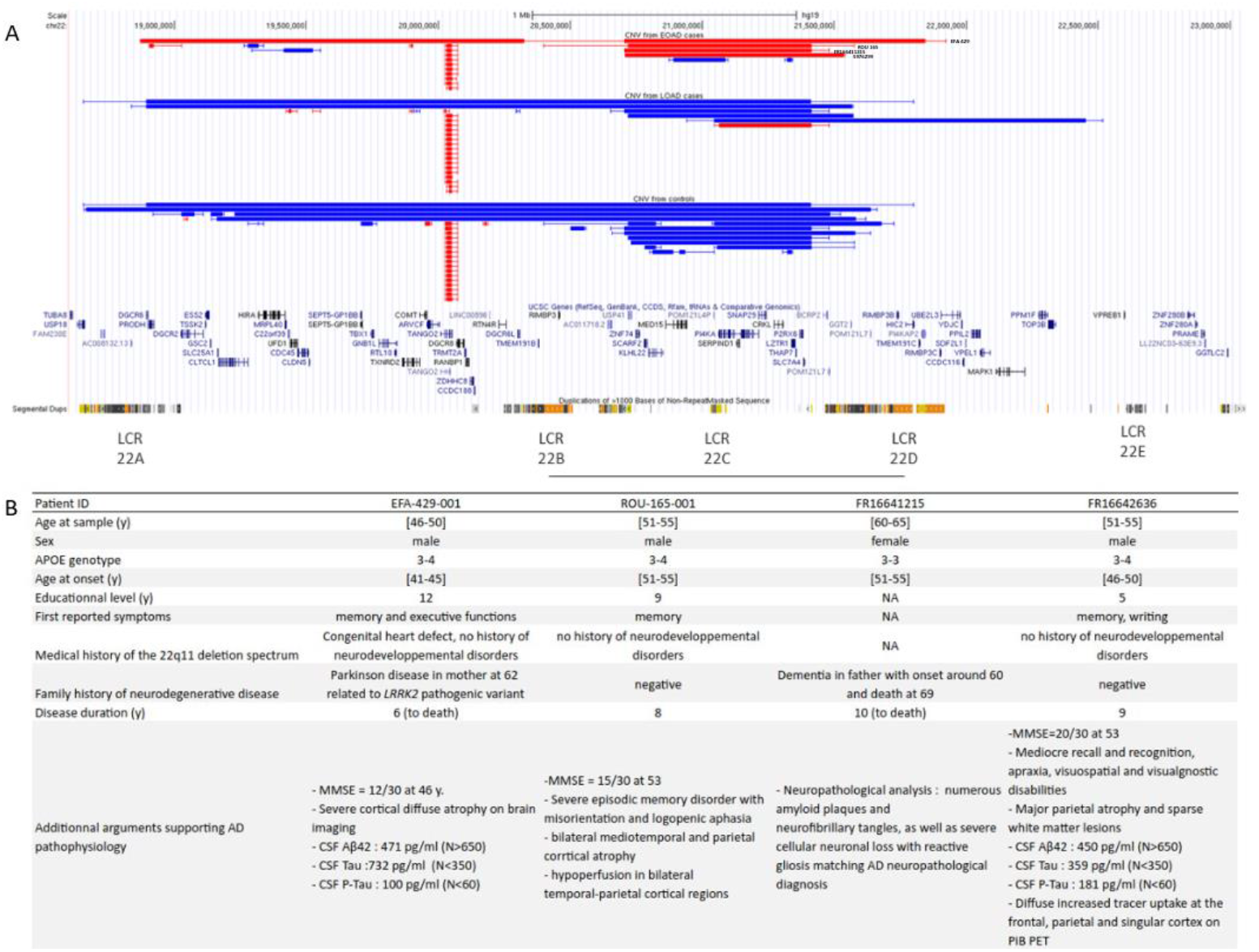
Focus on the 22q11.21 locus. **A. UCSC genome browser view of CNVs identified in the main analysis** In red: deletions; In blue: duplications. Large areas represent coordinates of CNVs as detected by CANOES. Thinner areas show breakpoints uncertainties, i.e. regions between two targets of the sample-specific capture kit. Genes are indicated below, and the regions of segmental duplications appear at the bottom part. Low copy repeats (LCR) underlying recurrent rearrangements by non-allelic homologous recombination (NAHR) are indicated below. A black line underlines the locus prioritized here. Note that not all duplications are taken into account in the statistical analysis, only those encompassing a whole transcript are included. **B. Clinical information of carriers of the deletion in the main analysis** Y: years; AD: Alzheimer Disease; EOAD: Early Onset AD; LOAD: Late Onset AD; MMSE: Mini Mental State Examination; CSF: Cerebrospinal Fluid

A total of 4,989 transcripts of 1,716 genes fulfilled our criteria for dosage association analysis,i.e. belonging to non-frequently deleted or duplicated genes, showing at least 4 rare CNVs in the dataset, including at least one (partial or complete) deletion and one complete duplication.

Of them, dosage analysis comparing EOAD patients to unaffected controls prioritized 44 transcripts (FDR≤10%, Table 3) from 18 genes, at five *loci*, as 13 genes mapped to chromosome 22q11.21 (0.0271<FDR<0.0386), two other contiguous genes mapped to 19q13.32 (*ERCC2*-*KLC3* locus, 0.0271<FDR<0.0412), and three genes were excluded (Table 3, supplementary information).

Three single-gene loci showed an FDR≤10%, namely *MBL2* (FDR=0.0271), *FADS6* (FDR=0.0271) and *ADI1* (FDR=0.0916). The *MBL2* signal did not remain after adjusting for population structure and same adjustment decreased the significance for *FADS6* while it did not affect the other loci (Figure S14, Figure S15, Table S11). The *ERCC2*-*KLC3* signal was driven by complete duplications (Table S9) and we showed that these rare duplications occurred on an *APOE4*-linked haplotype (Fisher’s exact test p-value=4.26×10^-13^) on chr19q13.32 (TableS 12, S10, Figure S3). The *ERCC2*-*KLC3* association signal did not remain significant after adjusting for *APOE*-ε4 dosage (OR=1.58 [0.85; 2.99], p=0.148), while other loci remained unaffected (Table S10).

The 22q11.21 region is known for rare recurrent deletions and duplications driven by non-allelic homologous recombination (NAHR) through low copy repeats (LCR) labelled LCR 22A to 22E (Figure 2). The region where all transcripts with FDR≤10% clustered is located between LCR 22B and 22D (labelled as the “central region”), which do not encompass the critical region for DiGeorge (velo-cardio-facial) syndrome (22A-22B). We observed 22q11.21 central region deletions in four EOAD cases (0.1%), and this region was also prioritized in the deletion-only analysis (Table S8; 0 control carrier, 0 to 1 (0.01%) LOAD carrier depending on the CNV coordinates). One of the EOAD cases carried a larger LCR 22A-22D deletion, typical of DiGeorge syndrome (Figure 2). He had no specific neurodevelopmental history, but he was born with congenital heart disease and presented first signs of cognitive decline at age 43, without any signs of Parkinsonism. The AD diagnosis was confirmed with CSF biomarkers (Figure 2). After targeted analysis in both unaffected parents, we showed that the deletion occurred *de novo*. Interestingly, duplications mirroring the LCR 22A-22D or the 22B-22D deletions, or affecting the 22C-22D or 22C to 22E regions, were observed in 8 to 10 controls (0.08 to 0.1%, depending on the CNV coordinates), in 4 to 5 LOAD cases (0.05 to 0.06% Figure 2), and in 0 to 1 EOAD case (0 to 0.02%) (Table S13). Overall, this suggests a dosage effect with deletions increasing AD risk and duplications decreasing the risk or delaying ages at onset. Performing the dosage analysis at the level of the whole cohort including LOAD patients using an ordinal regression, the association signal remained nominally significant (OR=0.16 [0.04 ;0.45] (p=3.5×10^-4^) (Table S7) as LOAD patients showed frequencies of deletions and duplications in between EOAD cases and controls.

Overall, DNA from 27 carriers (12 *ERCC2*-*KLC3*, two 22q11.2, one *MBL2*, three *ADI1* and nine *FADS6*) was available and CNVs were confirmed in 26/27 (one *FADS6* false positive (Table S14) - after removing the latter one, EOAD versus controls dosage analysis remained similar: OR=0.20 [0.07; 0.49], p-value=4×10^-4^), in addition to two 22q11.21 deletions confirmed in independent chip data. This further supports the accuracy of the calling method, which is consistent with our previous results of validation based on a large number of CNVs and comparing exome-DNA chip pairs^23^.

### Integrated loss-of-function analysis at the prioritized loci

Then, we performed a case-control integrated LOF analysis gathering CNV-deletions (complete and partial) with short truncating variants (nonsense, frameshift, canonical splice site SNVs/indels) among the genes belonging to the prioritized loci in the dosage analysis (Tables S6, S7). Although this integrated LOF analysis did not substantially modify the interpretation of the dosage analysis results, it narrowed the signal to the *SCARF2-MED15-KLHL22 locus* in the 22q11.21 central region. Indeed, we observed one LOF short variant in *SCARF2* in an EOAD case and 2 in *MED15* in 2 LOAD cases, whereas no control carried such LOF variants, as for CNV-deletions, and all the other genes at this locus appeared as less significant now.

### Replication of suggestive loci in large external cohorts

We aimed at replicating the *SCARF2-MED15-KLHL22* locus dosage, *ADI1* duplications and *FADS6* deletions cases enrichment, using chip data from 29,763 AD cases (3411 EOAD) and 359,928 unaffected controls, and exomes from African American AD cases (n=1462 including 182 EOAD cases) and unaffected controls (n=2393). As the *FADS6* signal was driven by a small recurrent deletion in the discovery (<10kb, Figure S6), which is not detectable in DNA chip data, we were not able to perform an unbiased mega-analysis. Regarding *ADI1*, replication results were in the same direction as the discovery analysis (Figure 3, Table S15) but significance was not achieved (Tables S15, S16). However, we succeeded in replicating the *SCARF2*-*KLHL22*-*MED15* gene dosage association at the 22q11.21 locus, as ordinal regression confirmed an EOAD<LOAD<control association (Tables S15, S16 Figure 3, replication OR_SCARF2_=0.40 [0.23;0.66], p=1.50×10^-4^), with a mega-analysis OR_SCARF2_=0.34 [0.21;0.53] (p=5.52×10^-7^), achieving both study-wide and exome-wide significance thresholds (OR to be interpreted per additional gene copy).

**Figure 3.**
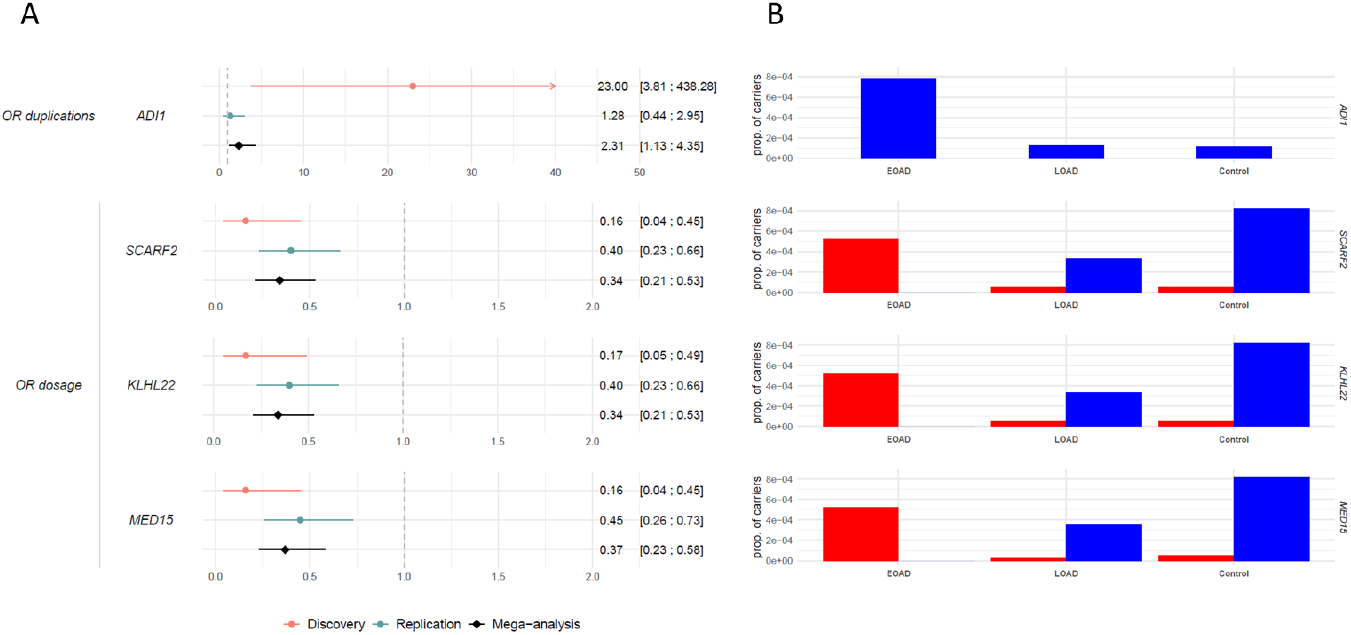
Mega-analysis of results from both main analysis and replication. **A. Forest plot associated with the mega-analysis of results from both discovery and replication**. Figure displays OR with 95% Confidence Intervals. For dosage analyses (*SCARF2, KLHL22, MED15*), OR should be interpreted for an increase of transcript copies by 1. i.e. a OR > 1 (resp. OR <1) means that duplications (resp. deletions) are associated with a higher disease risk (see Figure 1). OR: Odd-ratio **B. Proportion of deletions and duplications carriers by status and gene** In red: deletions; In blue: complete duplications. Data include both discovery and replication datasets.

### Assessing SCARF2 as a potential biological driver of the 22q11.21 association

Finally, we sought to provide an initial step toward understanding a potential biological basis for the *SCARF2*-*KLHL22*-*MED15* gene dosage association. We prioritized the *SCARF2* gene, as it belongs to the class F scavenger receptor family and is expressed in multiple cell types in the brain and in macrophages. Although a scavenger receptor role is yet to be demonstrated for SCARF2, such activity has been confirmed for other family members, including *SCARF1* and *MEGF10*, the latter even being a known receptor for Aβ^29, 30^. We therefore tested whether SCARF2 may influence Aβ cellular uptake by overexpressing *SCARF2* in HEK293 cells, and observed a significantly increased uptake of fluorescent Aβ42 peptides in SCARF2-transfected cells compared to mock and TDP-43-transfected cells, used as controls (Figure 4).

**Figure 4.**
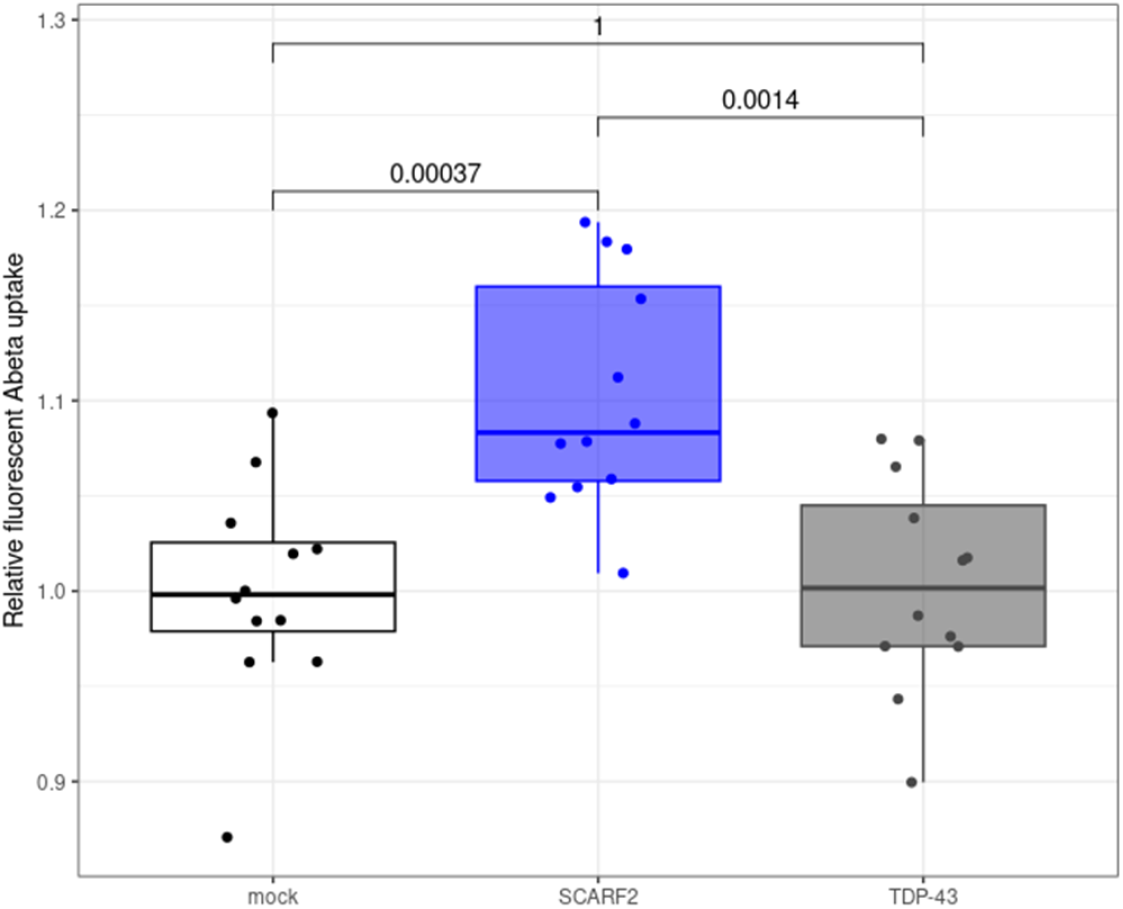
Aβ_42_ uptake in HEK293 cells. HEK293 cells transfected with an empty vector (mock) or overexpressing the SCARF2 or the TDP43 proteins were incubated with fluorescent Aβ_42_ peptides, and Aβ_42_ uptake was assessed by flow cytometry. SCARF2 overexpression was associated with a significantly increased uptake of Aβ_42_ peptides compared to the mock condition (p=0.00037), while TDP-43 overexpression did not affect Aβ_42_ uptake (p=1).

## DISCUSSION

Here, we identified an association with gene dosage at the central region of chr 22q11.2. Overall, the 22q11.21 locus showed enrichment of deletions in EOAD cases and mirroring duplications in controls, while LOAD cases remained in between. This region was not known to be associated with AD risk so far, despite a suggestive association in a family-based analysis^31^. In our gene dosage analysis, the association was exome-wide significant (p<2.75×10^-6^) and we are providing additional levels of evidence, with one of the patients with unaffected parents showing a *de novo* deletion, which is a strong genetic argument at the individual level, and a first biological argument on potential mechanism, as expression of *SCARF2* led to increased Aβ uptake.

Overall, central 22q11.21 deletions remain extremely rare. Here, no control carried such a deletion in the discovery, while duplications were enriched in unaffected controls. Notably, very few controls carrying central 22q11.21 deletions or larger ones (B-C or B-D) were identified in the replication (19/362,321, 0.005%), with a frequency that remains compatible with the absence of deletion in the discovery (0/9,559, i.e. frequency <0.01%). Among those having available information on parents in the UK biobank, 28.57% (2/7) had a parent with AD or related dementia, compared to 15.65% (35,569/227,256) in non-carriers of CNV in this region. However, rarity of deletion carriers prevented us to perform a powerful comparison (Fisher exact test p-value=0.3017). It should be noted that 10.36% of participants have an unknown status of family history in this database, and that such a deletion/duplication being driven by NAHR, they can be found as *de novo* events, hence limiting the interpretation of family history in this database for this locus. Beyond the fact that the penetrance of AD may not be full by the age of 65 in deletion carriers, most of those from the UK biobank are still rather young, so that some might still develop AD.

Interestingly, while our discovery was enriched in EOAD cases and highlighted at-risk deletions with mirroring duplications in controls, our replication was enriched in controls and highlighted the protective role of 22q11.21 central duplications with more power than the discovery alone. With an OR of 0.34 [0.18;0.56] in the mega-analysis, duplications appear as a strong AD risk-decreasing factor. So far, only a handful of genetic factors have been shown to significantly reduce AD risk, after exclusion of GWAS hits with ORs close to 1, namely the *APP* Icelandic variant A673T with a very strong effect (OR∼0.2), and three other AD risk-decreasing factors with ORs∼0.7: *APOE2*^32^, a rare missense variant in *PLCG2*^33^, and the very recent identification of a protective 19-bp deletion of an *APOE* enhancer in *APOE4*-positive African Americans, although carriers still show an increased AD risk compared to *APOE3* homozygozity^34^. Duplications of the 22q11.21 central region thus represent the second strongest AD-risk decreasing factor, after the A673T variant, which is both ultrarare and population specific, contrary to the 22q11.2, found in multiple ethnicities.

Of note, 22q11.21 central duplications reduce AD risk in the context of a complex determinism, conferring only relative “protection”. As one can identify AD risk factors in unaffected controls, we also identified a few duplications in LOAD duplication carriers. Of the four LOAD duplication carriers in the discovery, 2 (50%) carried at least one AD risk factor with OR>1.5, both being *APOE4* heterozygous and one of them also carrying a splice region variant in *ABCA7* (NM_019112.4:c.4764+5G>A), predicted to result in exon skipping and thus a probable AD risk factor with a moderate effect. As a comparison, 3/10 (30%) unaffected controls carrying a 22q11.21 duplication carried a similar AD risk factor (2 were *APOE4* heterozygous and one carried the *ATP8B4*:c.1183G>A, p.(Gly395Ser) classified as a modest risk factor with OR around 1.5)^35^, further demonstrating the complex nature of the genetic determinism of AD, and the need for integrated models gathering multiple risk and protective factors for an accurate AD risk prediction^36^.

In theory, two hypotheses might explain the association with gene dosage at the 22q11.21 central region. First, the association could be nonspecific to AD mechanisms but rather a modifier of AAO in individuals prone to develop AD. Indeed, carriers of the central 22q11.21 deletion have a higher risk of neurodevelopmental disorders^37^, as do the carriers of DiGeorge-syndrome associated deletions (22A-22B or larger) with higher penetrance, and lower average cognitive performances^38^. Whether or not it is clinically significant prior to AD onset, putatively reduced cognitive reserve might contribute to earlier AD onset ages^39, 40^. However, this hypothesis would require that duplication carriers have better average cognitive performances, which is not known, 22B-22D duplications being considered as variants of uncertain significance. Second, dosage of certain genes in this interval could be directly influencing the pathophysiology of AD. Given its presumed functions and expression, we hypothesized that *SCARF2* might act as a scavenger receptor for Aβ. We showed that overexpression of *SCARF2* led to increased Aβ uptake. Although these results will require further confirmation in other models and the underlying mechanisms remain to be determined, including a putative function as an actual scavenger receptor, they are consistent with the observed reduced risk in duplication carriers.

From a medical point of view, our results may be of interest for DiGeorge patients, among those with large deletions including *SCARF2*-*KLHL22*-*MED15* (22A-22C deletions or larger). DiGeorge syndrome presents with variable penetrance symptoms, including neurodevelopmental disorders ranging from mild intellectual disability, psychosis, and autism spectrum disorder to normal intelligence, congenital heart defects, palatal malformations, velopharyngeal incompetence, phosphocalcic metabolism disorders, immune deficiency, some morphological features and increased Parkinson disease risk^41^. However, not much is known about the aging of these patients, as most reports in adults involve individuals in their mid-30s on average and limited survival until late adulthood^42^. At least one patient (aged 56) showed AD-related neuropathology^41^, but neuropathological reports are scarce. Our results suggest that, as care of DiGeorge syndrome is improving, enabling patients to age, cognitive follow-up should be proposed, especially to those with a large deletion. This finding of a 22q11.21 association with AD is somehow reminiscent of the Trisomy-21 and *APP* duplication patients^8, 43, 44^. However, the impact of the latter may be much stronger, with full penetrance by the age of 65-70^11, 45^, probably because of the straightforward link between *APP* overexpression and Aβ production, while Aβ clearance involves multiple players, *SCARF2* now becoming a strong candidate Aβ scavenger.

Beyond the 22q11.21 locus, our study also highlighted CNVs in known AD genes directly contributing to the disease in carriers. Rare CNVs have long been implicated in diverse Mendelian disorders, including neurodegenerative diseases, in which they nevertheless remain an infrequent mechanism. For instance, only two *APP* triplications have been reported so far^43, 46^ while *APP* duplications account for ∼9% autosomal dominant EOAD families, and *MAPT* duplications represent a very rare cause of AD-mimicking primary tauopathy^26^, as found in some patients here. In line with Mendelian dementia genes, we show that rare CNVs also contribute to non-monogenic AD risk, as for *ABCA1* and *ABCA7*, where deletions represented a small proportion (<10%) of LOF alleles. Interestingly, we also identified a suggestive association of *CSTB* LOF with AD risk. *CTSB*, a GWAS-defined gene, encodes the Cathepsin B, an anti-amyloidogenic enzyme with Aβ peptide cleavage activity^47^, which is consistent with LOF alleles in cases and duplications in controls (see also supplementary information).

Here, we used a large existing dataset of exome sequencing data. We propose a novel QC and analysis strategy and obtained a dataset harmonized at the transcript level, to reduce both the recurrence issue at the CNV level and the heterogeneity among datasets. This allowed (i) the identification of high-quality CNV calls in known AD genes (Mendelian and risk factor genes) and (ii) identification of suggestive loci, despite the extreme rarity of the individual CNVs. This design was adapted to the switch from DNA chips to exomes, largely reducing the size of the smaller CNVs under analysis. Our study design was thus adapted to the identification of single gene association. Despite this design, the most significant finding was a multiple-gene recurrent CNV mediated by NAHR. The low number of single-gene association is probably related to a power issue in finding a burden of ultra-rare CNVs in a given gene, but our study design may be successful in finding such signals in future larger studies. In a replication attempt, we mostly relied on chip data that do not allow the calling of smaller CNVs, as the recurrent *FASD6* deletions. Genome sequencing may also be used, although at a high computational cost and with still limited sample sizes, especially for in terms of EOAD case data available^18, 19^. Novel long read sequencing technologies may also offer new opportunities, enabling the analysis of CNVs along with other complex variants^48, 49^, but sequencing costs remain high, limiting the sample sizes. In addition, we did not study the X chromosome, due to the complexity of the analysis models. Specific analysis of this chromosome may reveal potential variants associated with AD^50^.

In conclusion, we identified ultra-rare CNVs directly contributing to AD risk, including autosomal dominant CNVs, deletions in AD-associated *ABCA1* and *ABCA7* contributing to AD risk in non-monogenic AD, *CTSB* loss-of-function as candidate risk mechanism, and a small region at the 22q11.21 locus, deletions of which increase EOAD risk while duplications decrease AD risk. We are providing multiple layers of evidence, including a clear mirror/dosage signal with late-onset AD patients showing intermediate frequencies of deletions and duplications between EOAD and unaffected controls, a large replication with a multiethnic component, exome-wide statistical significance, one *de novo* deletion in an EOAD case, and a first mechanistic argument, all converging on the same interpretation. This result may be important for the follow up of some patients with DiGeorge syndrome who may present increased risk of AD. In addition, we identified the second strongest AD risk decreasing factor, 22q11.21 duplications. The *SCARF2* gene, located in the prioritized region at the chr22q11.21 locus, may be a therapeutic target, as overexpression increased Aβ uptake, hence most likely Aβ clearance, which is congruent with reduced AD risk in duplication carriers. While additional CNVs may contribute to AD risk, as suggested by burden CNV analyses at the exome and AD gene lists levels, such CNVs remain extremely rare events, limiting power.

## Data Availability

All data produced in the present study are available upon reasonable request to the authors

## ONLINE METHODS

Detailed methods are provided in supplementary information.

### Exome dataset, CNV Calling, quality control, and pathogenic CNVs identification

All samples were selected following the quality control detailed in ref.^3^, except pathogenic CNV carriers, who were not excluded from our starting dataset.

Only samples passing the quality control (QC) described in detail in ref.^3^ were selected for our analysis. Briefly, for the individual QC based on short sequence variants, all sequencing (Fastq) files were processed using the same BWA-GATK-based pipeline on the Cartesius supercomputer embedded in the Dutch national e-infrastructure. Samples with either a high level of variant missingness, a high suspicion of DNA contamination, a discordant genetic sex annotation, a non-European ancestry, a high number of novel variants (compared to dbSNP v150), a deviation from standard heterozygous/homozygous or transition/transversion ratios, and relatives up to the 3^rd^ degree were excluded. As cases, we considered all individuals with a diagnosis of definite or probable AD (using the NIAA or NINCDS-ADRA criteria depending on the date of diagnosis^51, 52^), based on clinical and paraclinical information including cerebrospinal fluid (CSF) AD biomarkers, when available. Individuals with unclear diagnosis were excluded (e.g., Braak stage I-II in cases, or Braak stages of V-VI in controls). In addition, pathogenic SNV and indel variant carriers in a list of Mendelian dementia genes were already excluded from stage-1 data in this dataset. We now applied the same analysis leading to the exclusion of 22 additional pathogenic variant carriers among cases and controls from stage-2 so that the megasample from ref.^3^ (stages 1+2) fulfils the same criteria (Table S1). Detailed methods for individual QC are available as a supplementary information file in ref.^3^. In order to homogenize EOAD/LOAD categorization across cohorts, we defined as EOAD, all cases with an age at onset (AAO) <66 years or age at last visit <66 if AAO is missing. Cases without numerical information for age were considered as LOAD.

Overall, our starting dataset contained 12,669 exomes from cases (including 4,150 EOAD) and 9,650 exomes from controls (Figure 1).

All participants provided informed written consent, as previously described^3^, and according to the declaration of Helsinki. This study, based on the retrospective analysis of existing data, was approved by the CERDE ethics committee from the Rouen University Hospital (CERNI/CERDE notifications 2017-015 and 2019-055). All the identifiers used in this document are known only to the laboratory members who addressed them.

### CNV calling

Bioinformatics tools detect CNVs from sequencing data through various approaches, including the relative distribution of read depth, relative positions of paired reads from each other, or multiple and partial alignments of reads^53, 54^. Such approaches applied to genome sequencing data show diverse recall and precision rates and require multiple combinations of tools, at high computational costs. When working from sequencing data obtained following capture, as for exome sequencing, using the read depth approach shows high levels of accuracy at the exome level for rare CNVs^20, 21^, not requiring combinations of tools. Exome-specific tools do not rely on intra-sample read depth variation along the sequence, but on region per region inter-sample comparison. We previously validated a CNV-calling workflow based on the CANOES tool^22^, which demonstrated good sensitivity (87.25 %) and positive predictive value (86.4-90.2%) for rare CNVs from exomes^23^. We adapted this workflow for a large dataset and applied it to BAM files. Samples with excessive calls were excluded, as were carriers of excessively large CNVs, which are unlikely germline, to avoid biases from clonal hematopoiesis associated with large blood-specific, age-related CNVs. We then assessed the presence of pathogenic Mendelian CNVs. CNV calling and quality control are detailed in supplementary information.

### CNV and transcripts filtering

For case-control analyses, we excluded CNVs cross-overlapping by ≥70% with common CNVs (frequency >1% in DGV-gold Standard and non gnomAD-non neuro non-Finnish Europeans, v2.1), CNVs with ≥50% overlap with segmental duplications, gonosomal CNVs, and those overlapping only one target (corresponding to monoexonic CNVs), to reduce false positives.

To address capture kit diversity, we worked at the transcript level (Figure S2). A transcript was considered as fully deleted or duplicated if all associated capture targets were affected in the respective capture kit. We then built a copy number matrix for each autosomal protein-coding transcript, with 5 possible states per sample and transcript post-filtration: (i) missing information, (ii) no CNV (copy number=2), (iii) complete or (iv) partial duplication, or (v) complete or partial deletion. We worked under the assumption that both complete and partial deletions likely result in haploinsufficiency. In dosage analyses, we converted states (ii), (iii) and (v) into a transcript copy number information.

Then, to avoid the analysis of rare CNVs affecting transcripts frequently deleted or duplicated, we built two sets of transcripts: (i) transcripts with <1% cumulative deletion frequency in public databases (set A, non-frequently deleted transcripts) and (ii) transcripts with <1% complete duplication frequency (set B, non-frequently duplicated transcripts).

### Statistical analyses

First, we performed burden CNV analyses, at the exome level, i.e. summing the total number of genes encompassed by a deletion or a full duplication (affecting coding transcripts, restricted to set A for the deletion analysis and to a set B for the duplication analysis). Then, we performed similar burden analysis, now focusing on two gene lists: (i) genes associated with AD in GWAS^2^ and (ii) genes having function in Aβ network^27^. Then, we focused on AD-associated genes by performing gene-based analyses of loss-of-function (LOF) variants including both deletion-CNVs and truncating SNVs/indels in genes previously associated with AD. Burden CNV analyses consisted in (i) EOAD-control association analyses using Firth’s logistic regression and (ii) ordinal regression using the whole dataset (EOAD, LOAD, controls),assuming an EOAD<LOAD<control effect (or conversely), as previously performed for burdens of rare SNV/indels at the gene level^3^.

Second, we performed a gene-level, exome-wide discovery analysis, consisting of a systematic gene-level association study of all protein-coding genes. This analysis was transcript based, to allow the putative identification of burdens of rare CNVs affecting only a proportion of transcripts of a given gene. Based on the stronger effect of rare short variants on EOAD risk and the expected extreme rarity of CNVs, limiting power, we compared EOAD cases to controls, and assessed gene dosage using complete/partial deletions (decreasing dosage) and complete duplications (increasing dosage), on the fusion of transcript sets A and B. Signals with FDR<10% were considered suggestive and were further assessed by ordinal regression using the whole dataset (EOAD, LOAD, controls).

Complementary analyses consisted in: (i) gene-level, transcript-based analyses on deletions only (all protein coding genes from set A), complete duplications only (all protein coding genes set B) and (ii) gene-based analysis of loss-of-function (LOF) variants (deletion-CNVs plus truncating SNVs/indels) in genes prioritized by the dosage analysis (integrated LOF analysis). In addition, we further performed adjusted analyses based on ancestry (Firth’s logistic model including the first ten principal components) and on *APOE4* allele dosage, as sensitivity analyses.

### CNV confirmation

CNVs of interest were all manually reviewed. Targeted validation of CNVs was performed when DNA was available, using either quantitative multiplex PCR of short fluorescent fragments (QMPSF) or droplet digital PCR (ddPCR).

### Replication

Loci with FDR<10% in the dosage discovery analysis, and remaining after adjustments and refinement by the integrated LOF analysis, were evaluated in a replication analysis using genotyping data from the European Alzheimer & Dementia Biobank (EADB), the UK biobank (UKBB), deCODE, GR@ACE and exomes of African Americans from ADSP. Detailed sample selection, calling and QC procedures are reported in the supplementary information. Due to heterogeneous numbers of EOAD cases in the replication cohorts, we pooled them into a single replication dataset. To leverage the full replication dataset in a single replication test, we performed an ordinal regression. Then, discovery and replication datasets were pooled in a mega-analysis. Nominal replication significance threshold was set at 0.05, study-wide significance was set at 2.91×10^-5^ corresponding to 1,717 gene-level tests fulfilling the above-mentioned criteria, out of 18,050 genes resulting from the fusion of sets A and B, with an exome-wide significance set at 2.77×10^-6^ (Bonferroni correction). Hits were considered as replicated if they reached both significance thresholds.

### *In vitro cellular assay*s

To investigate *SCARF2* involvement in Aβ uptake, we overexpressed the *SCARF2* cDNA in HEK293 cells and used, as controls, the *TARDP* cDNA (TDP-43 protein), which is not known to influence Aβ uptake or relate to Aβ-associated pathways, and a Mock control. Cells were then incubated with a pH-sensitive fluorescent Aβ conjugate (Aβ^pH^, Protonex GreenTM). Aβ internalization was quantified by flow cytometry through mean fluorescence intensity (MFI) per cell. Transfection efficiency was assessed by FLAG immunolabelling followed by flow cytometry analysis.

## Acknowledgements

We thank the ADSP study for providing exome data from cases and controls (ADSP umbrella NG00067.v3). ADSP data were prepared, archived, and distributed by the National Institute on Aging Alzheimer’s Disease Data Storage Site (NIAGADS) at the University of Pennsylvania (U24AG041689), funded by the National Institute on Aging. The ADSP umbrella contains data from the Alzheimer’s Disease Neuroimaging Initiative (ADNI) database (adni.loni.usc.edu). As such, the investigators within the ADNI contributed to the design and implementation of ADNI and/or provided data but did not participate in the analysis or writing of this report. A complete listing of ADNI investigators can be found at:

http://adni.loni.usc.edu/wp-content/uploads/how_to_apply/ADNI_Acknowledgement_List.pdf.

We thank the high-performance computing service at the University of Lille.

This research has been conducted using the UK Biobank Resource under application number 63653 (to G.Nicolas) and 40980 (to S. Jacquemont).

Ace Alzheimer Center Barcelona acknowledges the support Fundación bancaria “La Caixa”, Fundación ADEY, Fundación Echevarne and Grífols SA (GR@ACE project). Also, the Spanish National R+D+I and Instituto de Salud Carlos III (ISCIII) for grants PI19/01301, PI19/01240, P22/I01403 within the European Plan *FEDER “Una manera de hacer Europa”*, and grants PMP22/00022 within the plan *Proyectos de Investigación de Medicina Personalizada de Precisión*, within the European plan Next Generation EU. The support Joint Program for Neurodegenerative Diseases (JPND) towards PREADAPT project (grant NºAC19/00097) and AD-PriOMICS project (AC23/00038), and from German Federal Ministry of Education and Research (BMBF) for the DESCARTES project (BMBF grant Nº01EK2102B and 01EK2102A), German Research Foundation (DFG). IdR was supported by the ISCIII under the grant FI20/00215. Ace Alzheimer Center Barcelona research receives support from Roche, Janssen, Life Molecular Imaging, Araclon Biotech, Alkahest, Laboratorio de Análisis Echevarne, and IrsiCaixa. Some control samples and data from patients included in this study were provided in part by the National DNA Bank Carlos III (www.bancoadn.org, University of Salamanca, Spain) and Hospital Universitario Virgen de Valme (Sevilla, Spain).

Part of the work was carried out on the Cartesius supercomputer, which is embedded in the Dutch national e-infrastructure with the support of SURF Cooperative. Computing hours were granted to Henne Holstege by the Dutch Research Council (‘100plus’: project# vuh15226, 15318, 17232, 2020.030; ‘Role of VNTRs in AD’; project# 2022.028, ‘Alzheimer’s Genetics Hub’ project# 2022.031 and 2024:036).

## Supplementary material

Contains 16 supplementary tables, 18 supplementary figures, and a supplementary information document

## Data Availability

Summary statistics are available upon request. CNV counts (deletions and full duplications) in all genes are available as a supplement. Access to deidentified raw data of the ADES study can be requested using the Alzheimer Genetics Hub. Authors do not own data from ADSP and UK biobank, which can be requested through their respective websites following specific procedures. CNV calling data have been deposited on the NIAGADS portal.

Dataset page: https://dss.niagads.org/datasets/ng00176/

Study page: https://dss.niagads.org/studies/sa000081/

