## supplementary text and figures for "Exome analysis of 22,319 individuals links extremely rare CNVs and 22q11.21 dosage to Alzheimer’s risk"

### SUPPLEMENTARY INFORMATION

#### INDEX

### DETAILED METHODS

#### Exome data

##### *Cohorts*

**ADES-FR (Alzheimer Disease European Sequencing-France, France):** WES of 1,731 cases (1,413 EOAD, 318 LOAD) and 1,017 controls recruited either at the French national reference center for young Alzheimer Disease at Rouen (CNRMAJ-Rouen; PHRC-GMAJ and ECASCAD study), or through the European Alzheimer's Disease Initiative (EADI) consortium[1] or the French exome project (FREX).

**AgeCoDe-UKBonn (Aging, Cognition and Dementia in primary care, Germany):** WES of 371 cases (99 EOAD, 272 LOAD) and 1 control from the combination of the German study "Aging, Cognition, and dementia" (AgeCoDe)[2] and patients' recruitment at the interdisciplinary Memory Clinic of the department of psychiatry and department of Neurology at the University Hospital in Bonn (UKBonn). The control individual and all LOAD patients are from the AgeCoDe study and all had ≥ 75 years of age (AAO for cases, current age for control). EOAD cases were recruited through UKBonn cohort.

**Barcelona-SPIN (Sant Pau Initiative on Neurodegeneration, Spain):** WES of 50 EOAD patients and 9 controls recruited through the multimodal Sant Pau Initiative on Neurodegeneration (SPIN) cohort (https://santpaumemoryunit.com/our-research/spin-cohort/)[3] and for which neuropathological samples were obtained from the Neurological Tissue Bank of Biobanc-HospitalClinic-IDIBAPS.

**AC-EMC (Alzheimer center - Erasmus University Medical Center, The Netherlands):** WES of 108 cases (86 EOAD, 22 LOAD) from the Alzheimer center Erasmus MC cohort (AC-EMC) that are patients referred to the Department of Neurology of the Erasmus Medical center of Rotterdam.

**ERF (Erasmus Rucphen Family, The Netherlands):** WES of 4 cases (1 EOAD, 3 LOAD) and 319 controls from the Erasmus Rucphen Family (ERF) study, a family-based cohort study that is embedded in the Genetic Research in Isolated Populations (GRIP) program in the Southwest of the Netherlands.

**Rotterdam study (The Netherlands):** WES of 366 cases (3 EOAD, 363 LOAD) and 1,514 controls recruited in the prospective population-based cohort from Rotterdam study[4], focused on chronic disabling conditions of the elderly[5].

**ADC-Amsterdam (Amsterdam medical center, The Netherlands):** WES of 762 cases (502 EOAD, 260 LOAD) and 303 controls from patients who visit the memory clinic of the Alzheimer center at the Amsterdam University medical center[6]. Controls are individuals diagnosed with psychiatric and subjective cognitive complaints.

**Netherlands Brain Bank (The Netherlands):** WES of 169 cases (51 EOAD, 118 LOAD) and 52 controls obtained from DNA isolated from brain tissues.

**100-plus Study (The Netherlands):** WES of 64 LOAD patients (AAO ≥ 100 years) and 282 controls ≥ 100 years of age from the prospective cohort study of cognitively healthy centenarians[7].

**EMIF-AD 90-plus Study (European Medical Information Framework for Alzheimer's Disease, The Netherlands):** WES of 69 controls ≥ 90 years of age[8].

**CBC (Control Brain consortium, UK):** WES of 111 cases (33 EOAD, 78 LOAD) and 250 controls derived from brain banks in UK and USA[9].

**PERADES (Defining Genetic, Polygenic and Environmental Risk for Alzheimer's Disease using multiple powerful cohorts, focused Epigenetics and Stem cell metabolomics UK):** WES of 3,450 cases (1,110 EOAD, 2,340 LOAD) and 609 controls recruited across UK, Italy and Spain for the "Defining Genetic, Polygenic and Environmental Risk for AD" study.

**ADSP (American Alzheimer Disease Sequencing Project, USA):** WES of 5,502 cases (817 EOAD, 4,685 LOAD) and 5,228 controls from the ADSP Discovery phase (stage 1 in ref.[10]) and the ADSP Discovery extension and Augmentation phase (stage 2 in ref.[10]). Of note, controls from the Discovery phase were all ≥ 60 years old and were selected as those having the least probability of converting to AD by age 85[11].

##### *BAM files sources*

BAM files were retrieved either from the pipeline described in ref.[10] or directly downloaded from the dbGAP website (ADSP-stage 2, 1554 samples). For most of the exomes from ref.[10], BAM files were retrieved from the original processing pipeline after quality control. BAM were aligned on the GRCh37 version of the human genome. BAM from the ADSP Discovery extension and Augmentation phase were downloaded directly from the database of Genotypes and Phenotypes (dbGAP) website (N=1588). Of note, ADSP-stage 2 BAM files were aligned to the GRCh38 version of the human genome, whereas other BAM files were aligned on the GRCh37 version. As our analysis is based on transcripts affected by CNVs and because reference genome was not specific to cases or to controls, we expect that the different reference genomes do not affect our analysis. CNV calling and annotations were made for each sample according to the genome version. Then, data were merged to be analyzed jointly only at the transcript level.

All samples were processed on either the Cartesius supercomputer or the CEREBRO cluster from the sequencing facility of the University of Rouen Normandie (ASGARD platform).

#### CNV calling and quality control

We applied a workflow centered on the CANOES tool, which is based on the distribution of the depth of coverage information across samples. It includes a correction based on GC content of each exome sequencing capture kit-specific target to reduce the background variability often observed in NGS data[12].

We chose the use of CANOES because of its high performance based on a prior validation study[13]. Of note, as CANOES (and other read-depth comparison tools adapted to sequencing data obtained following capture) is based on the comparison of read depth on a specific target to a matrix computed from samples from a same batch, it is associated with better performances in detecting rare CNVs than common ones. Indeed, a very common variant (several dozens of percent in frequency) would likely be missed because of natural variability in each batch. Even if CANOES can detect some common variants, among those with a rather low frequency (e.g. 1-10%), its performances have not been assessed precisely on such variants. Because rare CNVs are more likely to be associated with higher AD risk, as for short variants, and because of the performances of read-depth comparison tools enabling CNV detection from exome sequencing data, we focused on rare CNVs (frequency <1%, see below for filtering), with two sets of CNVs considered: first, to detect pathogenic CNVs in Mendelian genes, we favored sensitivity over specificity and included all rare CNVs, second, for case-control analyses, we focused on rare CNVs affecting at least two targets to reduce putatively false calls.

Indeed, in our validation study[13], the rate of false positive calls among CNVs overlapping one target reached 30% as compared to 9.8% in CNVs called by ≥ 2 targets.

##### *CNV calling using a CANOES-centered workflow and optimization of CANOES source code for large dataset*

For each capture kit, targets covering the same exon and separated by less than 30 bp were merged. Then, for each sample, the number of reads covering each merged target was determined using BEDTools[14]. For each CNV-calling batch, we removed non-informative regions, namely regions where >90% of the samples showed less than 10 reads on the target. Finally, CANOES was run on each CNV-calling batch to generate CNV calls.

Because our working dataset is built from multiple studies, and to ensure a good accuracy of CANOES[15], we focused on samples as homogeneous as possible in batches of at least 50 individuals (Table S2). Samples prepared with a same capture kit from a same study were grouped primarily by sequencing batches, when information was available, otherwise by sequencing center else study. However, the latter option led to subsets of samples that were too large for an optimal use of CANOES as the time needed for processing an individual is proportional to the size of the input dataset.

To improve performances (computational time and resources needed) of CANOES on large datasets including ≥ 100 individuals, we slightly modified the original version of CANOES. Indeed, during the process, CANOES needs to generate an NxN matrix based on read counts similarity between samples. In the original version, CANOES may be executed sequentially or in parallel over a list of samples. In both cases, the NxN matrix was newly generated before the CNV calling for each sample, despite its consistency across all processed samples. If the time needed to completely analyze (including the matrix generation and the CNV calling) is less than 14 minutes for a dataset of 100 samples, it can go up to more than 11 hours for large datasets including 1000 samples. In this updated version of CANOES, we separated the matrix generation from the calling process, such that the NxN matrix is generated only once for an input dataset. Then, the calling process may be executed in parallel over all samples. In a comparative study of the two CANOES versions in a similar setting of computational resources and parallelization (192 Gb of RAM and 32 threads), we observed that the modification helped us to save ~2 minutes and ~9 hours for datasets with respectively ~100 and ~1000 samples (Figure S12).

##### *Quality Control and detection of large likely mosaic CNVs*

After CNV calling, we excluded all individuals with ≥50 calls, indicating an excess of variability in the sample's read distribution compared to other samples from the same CNV-calling batch and following CANOES user instructions[15].

Then, to account for potential biases due to clonal hematopoiesis associated with large mosaic, blood-specific age-related CNVs, we excluded carriers of CNVs that are too large to be likely germline CNVs. Due to its high sensibility, CANOES is indeed able to call mosaic events appearing in > 30% of cells[16], although it is not possible to sort CNVs that are likely germline from the ones that are likely not, based on CNV calling information. Such mosaic CNVs could be acquired early by mutation during the development process, or later in case of blood-specific, age-related events[17]. Both are associated with a lower frequency of cells carrying the mutation leading to a weak signal. Combined with the fact that they encompass low complexity regions, their calling may be less accurate and such large CNVs can be called as multiple, smaller CNVs. Thus, for each sample and each chromosome, we computed the cumulative size of detected CNVs of the same type (deletion/duplication). For each cumulative size per chromosome greater than 2.5Mb encompassing a range of 10Mb, we proceeded to a manual visualization (Figure S1) on the UCSC genome browser. Literature review was performed to support decision making. Carriers of candidate large blood-specific, age-related CNVs were excluded from the analysis (Table S3).

In addition, 6 samples showed a suspected overamplification of several genes related to a custom capture kit, and were further excluded from the analysis.

##### *Pathogenic CNV detection*

For pathogenic CNV interpretation, we considered partial deletions of *PSEN1* involving exclusively exon 9 (NM_000021.4) or both exons 9 and 10[18,19], complete duplications/triplications of *APP*[20], *MAPT*[16], *SNCA*[21,22] as well as any coding deletion of *GRN*[23] as causing either AD or a differential diagnosis and excluded carriers of those for further analyses.

##### *CNV filtering*

###### *Frequency*

For each CNV, we computed two frequencies by considering independently information from two public databases: the Database of Genomic Variants (DGV) gold Standard section[24] and the non-neuro non-Finnish European section of the gnomAD database v2.1[25]. For each CNV and each public dataset, corresponding frequency was computed as follows:

$$\text{frequency} = \frac{n_{\text{carriers}}}{n_{\text{total}}}$$

where $n_{carriers}$ corresponds to the number of carriers of a given CNV (of the same type [deletion or duplication]) overlapping mutually ≥ 70% with the CNV of interest; $n_{total}$is the total number of individuals included in the study revealing the candidate CNV. In case of several candidate CNVs from several studies, all carriers of such CNVs were added together in the numerator $n_{carriers}$ whereas the denominator ($n_{total}$) corresponded to the highest sample size among all studies contributing to candidate CNVs in order to count individuals contributing to several candidate CNVs only once. Besides this may lead to an overestimation of the frequency, it makes our definition of “rare CNV” more stringent.

All CNVs with a frequency >1% in at least one of the above-mentioned public databases were excluded from the case-control analyses.

###### *Segmental duplications*

Segmental duplication regions were extracted from the UCSC Table Browser (https://genome-euro.ucsc.edu/cgi-bin/hgTables). For each CNV, we computed the proportion of bases overlapping segmental duplication regions. All CNV with a ≥50% overlap with segmental duplication regions were excluded from the analyses, as they are considered as highly variable across individuals and hardly callable.

#### Information at the transcript level

To avoid potential bias due to heterogeneity of capture kits, we worked at the transcript level. Indeed, classic CNV annotation tools, which are based on transcript coordinates defined by public databases (e.g., RefSeq), may not be applied to heterogeneous datasets, as the definition of a complete versus partial gene duplication or deletion usually relies on the actual definition of a given gene, based on genome coordinates. As some capture kits vary on the regions actually targeted for a given gene (e.g. capture of untranslated regions or not, capture of all coding exons from all known transcripts or not), this may lead to heterogeneous definitions of partial or complete deletions/duplications, some transcripts being falsely reported as partially affected despite they should be considered as full events (see for a theoretical example Figure S2). Thus, we built our own annotation pipeline that relied on each individual's capture kit to define whether a given CNV partially or completely affected a given transcript. We redefined the transcripts positions according to every capture kit used (Figure S2). A transcript was considered as completely deleted or duplicated if all targets linked to this transcript were affected by the event. For all comparisons between capture kits, CNV calls and public databases, we used a combination of the BEDTools suite[14] and homemade scripts.

This allowed us (i) to adapt the number of analyzable individuals for each transcript, (ii) to more easily highlight transcripts of interest affected by multiples CNVs with different coordinates, and (iii) to exclude transcripts affected by a recurrent small transcript-specific CNV without excluding the other transcripts of the same gene that were not affected by it. For each deletion and each duplication, we reported if it encompasses partially or completely a transcript.

Based on this classification and on calls statistics, we built a copy number matrix per autosomal protein-coding transcript, to harmonize heterogeneous data at the transcript level and to determine the missingness and hence which transcripts were eligible for case-control analysis. For each sample and each transcript remaining after filtration, five possible states were determined: (i) missing information, i.e. the corresponding capture kit does not target the transcript or the region was not covered enough (≤10 reads) for 90% of the samples and no CNV encompassing this transcript has been detected from targets on neighboring genes or (ii) no CNV overlaps the transcript [copy number = 2] or (iii) a complete or (iv) a partial duplication overlaps this transcript or (v) a complete or a partial deletion overlaps this transcript. In our five-state categorization, we worked under the assumption that complete and partial deletions likely result in haploinsufficiency. To explore the dosage effect, states (ii), (iii) and (v) were converted into a transcript copy number information.

Then, to avoid the analysis of rare CNVs affecting transcripts frequently deleted or duplicated, we built two sets of transcript, based on Refseq protein-coding transcripts (assessed, 26/10/2022): (i) a set of transcripts with a cumulative frequency of deletions (partial or complete) of less than 1% in public databases (set A, corresponding to non-frequently deleted transcripts) and (ii) a set of transcripts with a cumulative frequency of complete duplications of less than 1% in public databases (set B, corresponding to non-frequently duplicated transcripts), based on the above-mentioned databases. Of 61,092 Refseq autosomal transcripts (related to 18,050 genes) targeted by at least one capture kit used in this study dataset, set A contains 57,489 non-frequently deleted transcripts (related to 17,227 genes), set B contains 60,108 non-frequently duplicated transcripts (17,846 genes) and the fusion of sets A and B contains 60,503 transcripts (18,050 genes).

A catalog of genes with partial/complete deletions and duplications and frequencies among EOAD cases, LOAD cases, and controls is provided as extended dataset.

#### Analyses and statistical tests

For each analysis, we compared EOAD cases versus controls using logistic Firth's regressions because of the rarity of CNVs and then, EOAD, LOAD, Controls ordinal regression analyses, to take advantage of the whole dataset.

All statistical analyses were performed using R version 3.6. Firth's logistic regressions were performed using the "logistf" function from "logistf" package[26]. All ordinal regressions were perfomed using the "clm" function of the "ordinal" package.

**Burden CNV analyses**

We performed burden CNV analyses, first at the exome level (all coding transcripts, restricted to the set A of transcripts for the deletion analysis and to the set B of transcripts for the duplication analysis), and then using two gene lists. To overcome the variability in sequencing techniques, we excluded all transcripts with a significant differential missingness between disease status ($p<1E-5$ in a $\chi 2$ test) from this analysis.

After the exome-wide levels restricted to set A or set B, we focused on two lists of genes: (i) genes associated with AD in GWAS[27] and (ii) genes having function in Aβ network[28] (and see below). For each analysis, deletions (partial and full) and duplications (full only) were analyzed separately. Models included either the information of the presence/absence of CNV in a gene included in the list of interest or the number of genes of the list encompassed by a CNV.

$$\text{logit}\left[\mathbb{P}(\text{case} \mid \text{CNV, list})\right] = \beta_0 + \beta_1 \times \mathbb{1}_{(\text{presence of a CNV encompassing genes in the list})}$$

$$\text{logit}\left[\mathbb{P}(\text{case} \mid \text{CNV, list})\right] = \beta_0 + \beta_1 \times \text{number of genes in the list being encompassed by a CNV}$$

**CNV-dosage analysis**

To perform an EOAD cases vs controls transcript-based association study on protein-coding genes using a dosage strategy as the primary endpoint, we worked on the fusion of transcript sets A and B and used rare partial and complete deletions and complete duplications. Dosage analyses were restricted to transcripts affected by a CNV in at least four carriers (including at least one deletion and one complete duplication) in our dataset and to the subset of individuals with available information relative to the transcript, i.e. individuals without missing information in the copy number matrix. This led us to perform 1,716 gene-level tests based on 4,989 transcripts.

For each transcript and each individual, the dosage information refers to the number of copies. Dosage equals 2 for individuals without any deletion nor full duplication affecting the transcript. Each duplication affecting the entire transcript increases the dosage by 1, whereas any deletion affecting partially or fully the transcript decreases the dosage by 1. Firth's logistic regression model is:

$$\text{logit}\left[\mathbb{P}(\text{case} \mid \text{dosage})\right] = \beta_0 + \beta_1 \times \text{dosage}$$

such that OR ($=\exp(\beta_1)$) associated with dosage information should be interpreted for an increase of transcript copies by 1, i.e. a OR > 1 (resp. OR <1) means that duplications (resp. deletions) are associated with a higher disease risk.

Associations were modeled *via* Firth's logistic regressions with status (EOAD *vs* controls) as dependent variable and dosage information (transcript copy number by individual) as independent variable. Based on a log-likelihood penalization, this method avoids infinite estimates due to separability problem that may arise in rare events analysis[29]. If the number of carriers of the different transcripts of a gene were similar, they were tested together. Otherwise, the transcripts were tested separately. Results with FDR<10% were considered as suggestive.

**Deletions/duplications analyses**

As a complementary analysis and to better understand signals from dosages, we performed transcript-based analyses in a deletions-only (complete and partial deletions, on set A) and in a complete duplications-only (on set B) setting. Regression was performed for each transcript from set A or B overlapping at least four deletions or four complete duplications in our dataset. For each transcript and each individual, the deletion (resp. full duplication) information refers to a binary variable: absence/presence of any deletions (resp. full duplication). Firth's logistic regression model is:

$$\text{logit}\left[\mathbb{P}(\text{case} \mid \text{presence of a CNV})\right] = \beta_0 + \beta_1 \times \mathbb{1}_{(\text{presence of a CNV})}$$

such that OR (=exp($\beta_1$)) associated with CNV (any deletion or full duplication) information should be interpreted for a CNV presence versus absence, i.e. a OR > 1 (resp.  OR <1) means that carrying a CNV is associated with a higher (resp. lower) disease risk.

**Joint CNV-deletions and loss-of-function indels/SNVs analyses**

At the gene level, we assessed the effect of all loss-of-function (LOF) variants in a list of AD risk genes and candidate genes. We included in this analysis all genes previously associated with AD in a GWAS study[27] and in our latest gene-based rare variants exome case-control study[10] (see below) as well as genes prioritized by the dosage EOAD – controls analysis (FDR≤10%).

LOF variants were defined as following:

- any deletion (partial or full) affecting at least one transcript within the gene. Only transcripts with less than 25% of missingness in the whole dataset were considered for this analysis;
- SNV/indel with the highest prediction of a LOF variant. To filter SNV/indels, we used the LOFTEE software[25] labeled as "High Confidence", as in ref.[10] (i.e., nonsense, canonical splice site variants and frameshift indels not affecting the last exon or the 50 last bp of the penultimate exon with no close rescue site as predicted by MaxEntScan software[30]). In addition, we performed an additional check on splicing variants, which can have diverse consequences that are more difficult to predict than nonsense variants and frameshift indels. We filtered out splicing variants affecting in-frame coding exons of less than 33 amino-acid residues (<100bp), as the likelihood of a LOF effect is lower. We also added manually the recurrent NM_019112.4:c.5570+5G>C *ABCA7* recurrent variant with a demonstrated effect on splicing and considered as a LOF variant[31]. We considered for the analysis all LoF-SNV with <25% of missingness, without differential missingness between cases and controls (based on a Fisher's exact test and p-value > $10^{-30}$) and a Variant Batch Detector (VBD) score <20[10] based on the list of individuals included in the current study.

Individuals with missing information in at least one transcript considered for deletion or in > 80% of gene position were not included in the analysis.

For each gene and each individual, the LOF information refers to a binary variable: absence/presence of a LOF variant as defined above. Firth's logistic regression model is:

$$\text{logit}\left[\mathbb{P}(\text{case} \mid \text{presence of a LOF})\right] = \beta_0 + \beta_1 \times \mathbb{1}_{(\text{presence of a LOF})}$$

such that OR (=exp($\beta_1$)) associated with LOF information should be interpreted for a LOF presence versus absence, i.e. a OR > 1 (resp.  OR <1) means that carrying a LOF is associated with a higher (resp. lower) disease risk.

**Ordinal regression analyses**

In parallel to the EOAD versus Controls analyses, we performed an EOAD, LOAD, Controls ordinal analyses to take advantage of the whole dataset, for each of the above-described sections, using the same covariates as Firth's model. This supposes a gradient effect with more deleterious variants in EOAD than LOAD than Controls and conversely for potential protective variants. P-values were estimated using a likelihood-ratio test.

**Establishing a list of AD-associated genes**

For the LOF analysis we focused on a list of AD-associated genes, in addition to genes prioritized at the dosage step. To build this list, we retained genes with the highest level of AD association, i.e. reaching genome-wide or exome-wide association, respectively for single variants and for gene-based tests. A recent large genome-wide association study (GWAS) reported 75 loci, most of them being associated through common non-coding single nucleotide polymorphisms (SNPs) with a modest effect on AD risk[27]. On the other hand, a burden of rare (frequency <1%) to ultra-rare (up to singleton) variants in *SORL1*, *TREM2*, *ABCA7*, *ABCA1* and *ATP8B4* as well as a handful of rare recurrent single variants demonstrated a wider diversity of effects, ranging from modest odds ratios for the least rare variants (OR=[1.5; 3], e.g., in *ABI3*, *TREM2* (R62H), *NCK2*, *SORT1*) to strong effects for some ultra-rare variants (e.g., loss-of-function *SORL1* variants)[10,31-35]. Of note, except for a few rare recurrent variants showing nominal association of their own, evidence for rare variant association with AD risk is generally obtained using burden tests gathering truncating variants with missense, predicted deleterious variants, at the gene level[36]. Burden tests performed on the category of truncating variants alone showed an exome-wide level of association for *SORL1* and *ABCA7* but only suggestive signals for *ABCA1* and *TREM2*[10]. For the latter genes, truncating variants remain extremely rare, explaining the difficulty to detect their association with AD risk due to insufficient statistical power in the current datasets. Importantly, genetic results are in line with known mechanisms, suggesting a deleterious effect of haploinsufficiency or loss of function of *SORL1*, *TREM2*, *ABCA7* and *ABCA1* on Amyloid β (Aβ) peptides aggregation. Likewise, extremely rare loss-of-function variants in the *TYROBP* gene encoding the main partner of *TREM2*, have recently been associated with AD risk[37]. Interestingly, some genes belong to both categories, i.e. genes mapping to GWAS loci with an AD association with a low OR (OR<1.5) and rare variants with a stronger OR (or even monogenic AD, as for *APP* itself), despite the absence of linkage disequilibrium between the two types of signals. Thus, GWAS results point to genes of interest that can show other types of genetic variants, without any genetic link, and with a much stronger individual effect. With the hypothesis that rare CNVs encompassing genes that have been previously associated with AD might be associated with a stronger risk with AD, we included in the AD gene list all genes mapping to genome-wide significant loci in the most recent study with the largest number of diagnosed AD cases[27]. With the hypothesis that rare CNVs may represent another type of gene deregulation that associate with a moderate to strong risk among genes associated to AD through burden test of rare variants at the gene level, we added to the GWAS list, the *SORL1*, *TREM2*, *ABCA7*, *ABCA1* and *ATP8B4* genes (most of them being already GWAS hits)[10] along with the recently identified *TYROBP* gene[37].

**Sensitivity analyses**

Sensitivity analyses include either adjustment for:

- ancestry based on the principal component analysis performed on the 1000G samples and described in ref.[10]. Firth's logistic model includes the first ten principal components and is thus:

$$\mathrm{logit}\left[\mathbb{P}(\mathrm{case} \mid \mathrm{X},\ \mathrm{ancestry})\right] = \beta_0 + \beta_1 \times \mathrm{X} + \alpha_1 \times \mathrm{PC}_1 + \ldots + \alpha_{10} \times \mathrm{PC}_{10}$$

- *APOE4* dosage being 0 for non-carriers of e4 allele, 1 for e4-heterozygote carriers and 2 for 4e-homozygote carriers. Firth's logistic model is thus:

$$\mathrm{logit}\left[\mathbb{P}(\mathrm{case} \mid \mathrm{X},\ APOE)\right] = \beta_0 + \beta_1 \times \mathrm{X} + \beta_2 \times APOE\ \mathrm{dosage}$$

The X variable represents either the dosage, the CNV or the LOF variant, and $\beta_1$ coefficient should be interpreted as described above according to the X variable.

#### CNV manual review and confirmation by an independent technique

The main analysis provided a list of prioritized transcripts/genes. Each transcript with an FDR below 10% was carefully checked in the UCSC genome browser showing all filtered and unfiltered CNVs separately at each transcript of interest, in cases and controls, along with the study and the capture kit information (Figures S3 to S7). This allowed us to detect signals driven by genes overlapping with repeats or duplicated gene in the genome despite CNVs not overlapping the 50% threshold set for repeats in the filtration steps, or genes for which the signal dosage analysis was driven by deletions whereas the transcript was not in set A, thus not relevant.

From each locus prioritized in the main analysis, we performed targeted validation of CNVs when DNA was available to us, using either ddPCR as previously described[38] or Quantitative Multiplex PCR of Short Fluorescent fragments (QMPSF). In patient EFA-429-001, we also confirmed parenthood using a set of informative microsatellites as the CNV that we detected was absent from parental samples.

#### Replication analysis

Loci of interest from the dosage analysis were assessed in a replication analysis based on genotyping data from the European Alzheimer & Dementia Biobank (EADB), the UK biobank (UKBB), Gr@ace, deCode and exome sequencing data from the ADSP-African American cohort.

CNVs from replication cohorts were annotated for frequency similarly to our primary analysis. Replication finally included the analysis of *ADI1* duplications as well as dosage analysis for the 22q11 *locus* (*SCARF2-MED15-KLHL22*). Indeed, the *FADS6* signal was partly linked to a recurrent small deletion in the discovery, the calling of which was not possible using DNA chip data. Because of the rarity of variants, replication consisted in performing the EOAD, LOAD, Controls ordinal analysis in the pooled replication dataset. Then, data from discovery and replication were pooled in a mega-analysis.

##### *EADB*

The European Alzheimer & Dementia Biobank dataset groups together AD cases and controls from 16 European countries (Austria, Belgium, Bulgaria, Czech Republic, Denmark, Finland, France, Germany, Greece, Italy, Portugal, Spain, Sweden, Switzerland, The Netherlands and the UK). The samples were genotyped in three different centers (Centre National de Recherche en Génomique Humaine, France, Erasmus Medical Center, Netherlands and Life & Brain, Germany) using the Illumina Infinium Global Screening Array. The sample and variant QC details can be found here[27]. From the EADB genotyping array data, we included 15,952 cases (2669 EOAD, 13,283 LOAD) and 20,335 controls not overlapping with ADES participants and after exclusion of relatives with a kinship >0.1 (supplementary information). CNV calling, QC and filtration procedures are described below.

CNVs were called using the standard settings of the PennCNV software. Measures of the B allele frequency (BAF) and log R ratio (LRR) from different genotyping centers were standardized. CNVs were considered if they were larger than 50 kb in size and covered at least 10 SNPs. 41735 samples passed initial QC.

To exclude doubletons between the EADB and the ADES that partially overlap, we performed relatedness estimations. We extracted the variants from ADES samples using the same QC criteria than those used in the primary analysis[1]. Then, (i) ambiguous variants (i.e. A/T or C/G), (ii) variants that are located in high LD regions as described here[39] as well as LCT (2q21), HLA and 2 inversion regions (8p23 and 17q21.31) and (iii) variants that could not be lifted to the GRCh38 assembly using Picard (https://broadinstitute.github.io/picard/) LiftoverVcf tool (v2.27.5) were excluded. Concerning the EADB dataset, we used all the available samples that passed the sample QC, excluding population outliers as described before[27], leading to the inclusion of 61,478 samples. Those samples were then imputed with the Trans-Omics for Precision Medicine reference panel – TOPMed[40,41]. Next, we only kept well imputed variants from the previous ADES list by excluding (i) variants showing an imputation quality $R2<0.8$ and (ii) variants having globally more than 10% of missingness by putting best guessed genotypes with a probability below 0.8 to missing. Finally, we performed a pruning of the remaining variants using plink v1.9[42] in the EADB dataset with an R2 threshold of linkage disequilibrium of 0.2 in a window of 500kb leading to the inclusion of 33,054 variants for the relatedness estimation of the EADB/ADES merged dataset (86,914 samples). The relatedness estimation was inferred by the GENESIS v2.32 package[43] to take into account population structure following the pipeline described here (https://bioconductor.org/packages/release/bioc/vignettes/GENESIS/inst/doc/pcair.html) and ref.[27] using 10 principal components for the PC-Relate step. Finally, 5114 EADB samples were removed using a kinship threshold of 0.09.

##### *UKBB*

The UK biobank is a large-scale database containing information on health, genetic data and lifestyle from almost 500,000 UK participants[44]. We selected Caucasian unrelated participants with available AD status (supplementary information). From the UKBB, we included 2050 cases (111 EOAD, 1939 LOAD) and 253,644 controls. CNV calling, QC and filtration procedures are below.

Among UKBB participants, we selected those without any kinship found (data-field 22021), from Caucasian ancestry (data-field 22006) and known AD status. Case-control status was defined based on ICD10-code F00 or G30 (data-field 41270). Age at onset was defined from date of birth and date of AD report (data-field 42020). Parents' status was based on family history reported by the participant and set to AD and related dementia, (ADD), no ADD or unknown (data-field 20107 and 20110).

CNVs called by both PennCNV[45] and QuantiSNP[46] were considered and merged using CNVision (https://martineaujeanlouis.github.io/MIND-GENESPARALLELCNV/). The following parameters were used for both algorithms: number of consecutive probes for CNV detection ≥3, CNV size ≥1Kb, likelihood scores ≥15. After these steps, an in-house algorithm based on CNV was applied to concatenate adjacent CNVs of the same type into one, according to the following criteria: a) gap between CNVs ≤150 kb; b) size of the CNVs ≥ 1000 bp; and c) number of probes ≥ 3. From the analysis, we excluded individuals with standard deviations of logRratio (resp. B allele frequency) ≥0.35 (resp. ≥ 0.08), a wave factor ≥ |0.05|, a call rate < 0.95 or being carrier of > 10 CNVs. We kept CNVs ≥50 Kb, covered by at least 10 probes and overlapping less than 50% with segmental duplication or HLA regions.

Finally, we included 2050 AD cases (among which 111 EOAD) and 253,644 controls. Parental dementia status was available for 89.64% of individuals.

##### *deCODE*

Approval for this study was obtained from the National Bioethics Committee and the Icelandic Data Protection Authority (VSN-19-129). Written informed consent was obtained from participants or their legal guardians prior to blood sample collection, and all sample identifiers were encrypted in accordance with the regulations of the Icelandic Data Protection Authority. The Icelandic Alzheimer's cohort (N=7193) has been previously described[31]. Briefly, a subset of patients was diagnosed with definite, probable, or possible Alzheimer's disease (AD) based on the NINCDS-ADRDA criteria[47]. For another subset, diagnoses were made according to the International Classification of Diseases, 10th Revision (ICD-10), using codes F00 or G30. Also, individuals who had been prescribed donepezil (Aricept) more than three times (N=461) were included in the case group, even when the aforementioned diagnostic criteria were not available. The case group was compared to population-based controls aged 55 years or older (N=77,432). Alzheimer's onset data were calculated from the first available Alzheimer's diagnoses or Aricept/Donepezil prescription. The mean age at onset was 80.6 years with standard deviation 7.7 of years. The sample was genotyped using Illumina: HumanHap, Omni and Infinium Global Screening SNP arrays as previously described[48]. BeadStudio (Illumina; version 2.0) was used to call genotypes, normalize signal intensity data and establish the log R ratio and B allele frequency at every SNP. Samples passing quality control were examined using PennCNV[45]. Additionally, results of analyses are provided after exclusion of individuals of artefact calls.

##### *GR@ACE*

The GR@ACE/DEGESCO cohort includes dementia patients and controls from Spain (N=20,080)[49,50]. The samples were genotyped using the Affymetrix Axiom SpainBA array. CNVs were called using the standard settings of the PennCNV software. Adjacent CNVs were merge (fraction 0.3). From the analysis, we excluded individuals with standard deviation of logRratio ≥0.38, B allele frequency drift ≥ 0.015, a wave factor ≥ |0.1|, a call rate < 0.95 or being carrier of > 40 CNVs. CNVs were considered if they were larger than 50 kb in size and covered at least 10 SNPs. CNVs that had >50% overlap with chromosomal centromeric and telomeric, or immunoglobulin regions plus 250kb were excluded. Individuals with low-quality SNP raw data, excess of heterozygosity, sex discrepancies and familial relations between samples (PI-HAT > 0.1875) were excluded from the analysis. A principal component analysis (PCA) was performed, and population outliers were removed (European cluster of 1000 Genomes). 15,866 unrelated samples passed QC including 7,348 AD cases and 8,518 controls.

##### *ADSP-AA*

The ADSP-African American cohort (ADSP-AA) is a subset of the ADSP cohort and was analyzed using the same pipeline as for the main analysis described in Figure 1. Patients and controls were selected based on the declaration of ethnicity reported in the data provided by ADSP.

#### Cellular assays

##### *Conjugation of pH-sensitive fluorescent Protonex Green™ with human Aβ (AβpH)*

The pH-sensitive fluorescent human Aβ conjugate with Protonex Green™ dye (AβpH) was prepared accordingly to (https://doi.org/10.1039/d1sc03486c) with minor modifications. First, 0.5 mg of human synthetic Aβ(1–42) (AnaSpec.Inc; #AS-20276) was monomerized with 100 µL HFIP at room temperature for at least 3 h before being aliquoted. HFIP was allowed to evaporate in the open tubes overnight in the fume hood and then dried down under high vacuum for 1 hour without heating. Dried peptides were stored at -20°C. Before use, monomeric Aβ(1-42) peptides were resuspended at 1 mg/mL in 1 M NaHCO3, pH 8.9. The amino-reactive Protonex Green 500-PEG12 SE dye (AAT Bioquest; #21219) was resuspended in DMSO to 10 mM concentration.

Then, Protonex dye and monomeric Aβ(1–42) were mixed at a 15:1 molar ratio (i.e, 66µL Protonex dye + 0.2 mg monomeric Aβ(1–42) in NaHCO3). The mixture was then diluted with ultrapure water (same volume as NaHCO3) to decrease viscosity and incubated at room temperature for 3 hours in the dark with rotating shaking to allow conjugation. The reaction mixture was then diluted with 1 mL ultrapure water and dialyzed using Amicon® Ultra-4 Centrifugal Filter Unit (Millipore) at 4,500 g for 45 minutes in a swinging bucket centrifuge to remove unbound dyes. The resulting concentrated solution was diluted again with 1 mL ultrapure water and dialyzed for 30 minutes. Then, the concentrated solution was diluted with 0.2 mL ultrapure water and lyophilized with a dry vacuum overnight to get the AβpH powder. Finally, this complex was monomerized using HFIP as previously described and resuspended at 500 µM in DMSO.

##### *Fluorescent Aβ42 uptake assay in SCARF2-expressing HEK293 cells*

Human Embryonic Kidney 293 (HEK293) cell line was grown in DMEM/F12 medium (Gibco/Thermo Fisher Scientific, Waltham, MA, USA), supplemented with 10% FCS (Eurobio, Les Ulis, France). Cells were seeded at $10^5$ cells per well on a 48-well plate 24 h prior to transfection, and transfected using the lipofectamine 3000 reagent (ThermoFischer Scientific) according to the manufacturer's protocol. The constructs included the pcDNA3 empty vector (mock), a pcDNA3 vector encoding human SCARF2 (NM_153334.6) with a C-terminal DYKDDDDK (FLAG) tag (#OHU28455D, Genscript, Piscataway, JW), and a pcDNA3 vector encoding the human TDP43 protein with a C-terminal FLAG tag. Forty-eight hours after transfection, cells were incubated for 2 hours with 200µL fresh medium containing 500 nM AβpH. Following incubation, cells were washed twice with PBS, detached using trypsin-EDTA solution, resuspended in FACS buffer consisting of PBS supplemented with 3% SVF, 1mM EDTA and 0.2 µg/mL DAPI (ThermoFisher, #62248), and then analyzed by flow cytometry.

##### *Assessment of transfection efficiency*

Transfection efficiency was controlled by Flag immunolabelling of SCARF2 and TDP43 in independent 48-wells, not treated with AβpH. Forty-eight hours after transfection, cells were detached using trypsin-EDTA and resuspended in PBS with 2% PFA and 0.2µg/mL DAPI. After a 15 min incubation, cells were centrifuged for 8 min at 750 × g at 4°C. Cells were washed with 200 µL of PBS-BSA 1%, then centrifuged again under the same conditions. Pelleted cells were resuspended in 100 µL 1X permeabilization buffer (eBioscience; #00-8333-56) containing anti-FLAG-L5-APC antibody (1/200, SONY; #3786535) and incubated for 30 min at 4°C. Subsequently, cells were washed with 900 µL of permeabilization buffer and centrifuged for 8 min at 750 × g at 4°C. Pelleted cells were resuspended in 300 µL of 1X permeabilization buffer and analyzed by flow cytometry, which indicated that at least 75% of HEK cells overexpressed the SCARF2 or TDP43 proteins (data not shown).

##### *Flow cytometry analysis*

Cells were analyzed by flow cytometry using a SONY ID7000 Spectral Analyzer and the data files were analyzed using the FlowJo software (V10, FlowJo, Ashland, OR, USA). A sequential gating strategy was applied to exclude dead cells (FSC-A vs DAPI), debris (FSC-A vs SSC-A), and doublets (FSC-A vs FSC-H). Outliers beyond the 2% isoline in contour plots were also excluded. Positivity thresholds for AβpH and FLAG-L5-APC were defined using dot plots of control populations (untreated and cells not overexpressing SCARF2). Geometric mean fluorescence intensities (gMFIs) were then calculated for each relevant population.

#### Code availability

All codes required for the analysis are available on the laboratory github at https://github.com/U1245/ExtremelyRareCNVContributingToADRisk.

Updated version of the CANOES pipeline is available at https://github.com/U1245/canoes-centered-workflow.

### ADDITIONAL RESULTS

*Dataset, CNV calling and QC*

We initially included 22,319 exomes (4,150 EOAD, 8,519 LOAD and 9,650 controls) (Figure 1). Following QC, we excluded 148 samples with excessive calls (≥ 50), 55 carriers of large likely somatic CNVs (26 controls, 28 LOAD cases, 1 EOAD case, Table S2, Table S3), and 6 samples with biased amplification due to a custom kit. There was no significant difference when comparing large likely somatic CNVs between all cases and controls and likely somatic CNVs were linked to age at last visit (used as a proxy for age at blood sampling) (logistic regression; Age: p-value=$6.78\times10^{-6}$; AD status: p-value=0.12). This suggests that such events are likely linked to clonal hematopoiesis and should thus be excluded from our germline CNV analysis study.

*CNV dosage analysis*

Of 61,092 Refseq autosomal transcripts (related to 18,050 genes) targeted by at least one capture kit used in this study dataset, set A contains 57,489 non-frequently deleted transcripts (related to 17,227 genes), of which 3299 (1040 genes) are affected by at least four deletions. Set B contains 60,108 non-frequently duplicated transcripts (17,846 genes), of which 3392 (1252 genes) affected by at least four complete duplications. The fusion of sets A and B contains 60,503 transcripts (18,050 genes), including 4989 transcripts (1716 genes) affected by at least four CNVs (including at least one deletion and one complete duplication). Results of the dosage analysis are described in the main document and Table 3.

*Sensitivity analysis*

In the sensitivity analysis, we adjusted our model for ancestry using the first ten principal components as covariates. (Figure S13 and Table S11). This adjustment did not affect ORs nor p-values in the dosage analysis except for two genes. First, variability associated with dosage in analysis of *FADS6* gene is higher after adjustment leading to 2-order of magnitude lower p-value. Second, in *MBL2* gene analysis, association became non-significant since the deletions observed were population dependent. Indeed, all carriers share a similar ancestry (Figure S14).

*Detailed results on the integrated LOF analysis at known AD-associated loci*

In *ABCA1*, we observed 3 carriers (all EOAD, AAO: 49, 51 and 55) of 3 distinct deletions, impacting exons 16 to 18, 32 to 34 and 47 to 50, respectively (Figure S7). The joint (EOAD vs controls) OR of *ABCA1* LOF variants was 5.77 [2.25; 17.06] (p=$2\times10^{-4}$), suggesting that *ABCA1* LOF is a moderate EOAD risk factor, while burden tests gathering LOF SNV/indels with missense variants showed more modest odds ratios[10]. Overall, *ABCA1* LOF variants remained extremely rare with only 13 EOAD (0.32%), 12 LOAD (0.14%) and 5 controls (0.05%) carriers. Deletions represented 10% (3/30) of the *ABCA1*-LOF alleles.

Deletions in *ABCA7* were observed in 4 EOAD cases (0.09%), 3 LOAD (0.04%) and 3 controls (0.03%). They represented 8.6% (10/115) of the *ABCA7*-LOF alleles, whereas a complete duplication was observed in one control (0.01%) and 2 LOAD cases (0.02%) (Figure S8). Deletions did not affect the order of magnitude of known AD or EOAD association of *ABCA7* with LOF variants in terms of OR, given the higher frequency of *ABCA7* LOF variants overall.

DNA was available for all 3 *ABCA1* and for 2 *ABCA7* deletion carriers and confirmed the existence of each deletion.

Our results are consistent with recent findings at the *TYROBP* locus[37] (OR (all AD versus controls) = 6.86 [0.73 ; 909.89], p= 0.1011), with 4 deletions in 4 LOAD cases, although there was no LOF SNV/indel, and deletions were not found in EOAD nor controls (precluding the use of ordinal regression)

Interestingly, CNVs covering *CTSB* and *APH1B* genes suggested a dosage effect with deletions of these genes observed in EOAD patients whereas duplications were observed in controls (Figure S10); although these genes did not appear in the top 10% FDR in the main dosage exome-wide analysis. Extending the analysis of these genes to LOAD, we observed 4 LOAD patients carrying a *CTSB* deletion and one LOAD duplication carrier (AAO=90) (Figure S11). There were no carriers of CNVs encompassing the *APH1B* gene among LOAD, and three controls carried a LOF SNV/indel. Thus, we performed an ordinal regression analysis assessing the dosage effect, considering a dosage similar to a deletion carrier for short truncating variant carriers (LoF variants and complete duplications). For *CTSB*, the ordinal analysis found an OR of 0.31 [0.15 ; 0.62], p = $8.05x10^{-4}$, further supporting a putative association of *CTSB* dosage with AD, whereas for *APH1B*, the OR was 0.45 [0.17 ; 1.16], p=$9.86x10^{-2}$.

Of note, LOF variants of *MAF*, *PLEKHA1* and *EPDR1* genes (including one deletion for each gene) were only detected in controls, leading to the hypothesis of a protective effect of LOF of these genes, which cannot be confirmed here as such events are extremely rare (Table S6, Figure S10).

### DETAILED DISCUSSION

*Rare duplications at the APOE locus*

In the protein-coding genome-wide analysis, one of the suggestive associations pointed to the *APOE* locus, which was not expected from a rare CNV analysis. However, this unexpected result, suggesting that rare duplications occurred on an *APOE4*-associated haplotype, further strengthen the validity of our global analysis. It is also a rare example of a rare recurrent CNV in linkage disequilibrium with a common GWAS locus, highlighting that this type of event can actually happen and should be considered in genome-wide analyses.

*Putative role of other genes at the 22q11.2 locus*

*MED15* is also expressed in the brain and is involved in inflammation and TGFβ and SMAD2/3 signal transduction, both implicated in AD pathophysiology[51], however with a less clear effect, and we did not prioritize this gene for cellular assays here.

*Suggestive association with FADS6 deletions*

Among other prioritized loci, *FADS6* deletions were associated with AD risk, although a small recurrent deletion, not detectable by DNA chips, was not present in one subpopulation from the discovery, thus requiring further replication (Figure S16). *FADS6* encodes the fatty acid desaturase 6, which is expressed in the brain. Its pathophysiological roles remain unclear, but *FADS6* expression is necessary for neuronal survival and neuroprotection against Tau phosphorylation after BACE1 silencing in cortical primary cultures[52].

*More CNVs might associated with AD risk*

In the analysis at the protein-coding genome level, we identified an enrichment of deletions and of duplications in cases. After focusing on the GWAS list of genes, the signal based on duplications trended to be reversed. This suggests that additional genes, not identified here, may play a role in AD determinism, and this highlights a limitation of analyzing CNVs as a whole, at the protein-coding genome or at gene-list levels, as increasing expression or decreasing expression of genes may show opposite effects on AD pathophysiology. For example, *APP*, *ABCA7* and *ABCA1* are all AD GWAS hits, but gathering their effect into a same analysis (if *APP* duplication carriers would not be excluded prior to case-control studies), would lead to non-interpretable results, as duplications of *APP* are expected to be enriched in cases but, conversely, deletions of *ABCA1* or *ABCA7* are expected to be enriched in cases as well. Similar reasoning can be applied to genes related to the Aβ network.

*CNVS add up to the signal on known AD genes from rare variants and identify novel probable independent associations from GWAS genes*

In this study, we identified and confirmed association of LOF alleles with AD in *ABCA7* and *ABCA1*. In *ABCA7*, the burden of LOF SNV/indels was already known to be associated with AD[31]. Joint analysis of CNVs and LOF SNV/indels did not modify the odds ratios much, remaining in the order of magnitude of 2 to 35[53], because (i) LOF alleles are not so rare (0.37% in controls; 0.86% in EOAD cases) and (ii) CNVs represented only 8.6% of all LOF alleles in the joint analysis. In *ABCA1*, where LOF variants are much rarer, we identified three distinct deletions, adding up to 27 LOF SNVs/indels. Deletions thus represent 10% of all LOF alleles. This analysis on *ABCA1* now allows us to consider LOF of *ABCA1* as a stronger risk factor for EOAD, compared to the known average odds ratios obtained by gathering LOF variants with missense, predicted damaging variants (OR=2.2 [1.6; 2.9] for LOF SNV/indels and missense variants with REVEL score >0.75 in ref.[10] and that also include missense variants with a demonstrated effect on AD risk, Tangier disease and HDL cholesterol deficiency[54,55], and OR=5.77 [2.25; 17.06] here in the joint LOF SNV/indels/CNV analysis. These results suggest that (i) some CNVs represent an extremely rare mechanism increasing the risk of AD but also that (ii) at the individual level, given the effect on AD risk of such LOF alleles, CNVs should not be ignored. Although risk variants are not used for genetic counseling, they may be used for the future of AD prevention in the context of precision medicine, along with other factors[56].

In addition, we assessed the presence of LOF alleles in the recently AD-associated gene *TYROBP*[37]. Four deletions were identified in LOAD cases, none in EOAD cases and none in controls. Three of the deletions likely shared similar breakpoints as a recurrent deletion described in Finland and also enriched in AD cases[37,57].

Methodological developments and perspectives

Here, we used a large existing dataset of exome sequencing data. We propose a novel QC and analysis strategy and obtained a dataset harmonized at the transcript level, to reduce both the recurrence issue at the CNV level and the heterogeneity among datasets. This allowed (i) the identification of high-quality CNV calls in known AD genes (Mendelian and risk factor genes) and (ii) identification of suggestive loci, despite the extreme rarity of the individual CNVs. Despite this design, the most significant finding was a multiple-gene recurrent CNV mediated by NAHR. This is probably related to a power issue in finding a burden of ultra-rare CNVs in a given gene, but this study design may be successful in finding such signals in future larger studies. In a replication attempt, we mostly relied on chip data that do not allow the calling of smaller CNVs. Whole genome sequencing (WGS) may also be used, although at a high computational cost and with still limited sample sizes, especially for EOAD cases. A recent analysis has been run on WGS of 6,646, mostly LOAD cases (average AAO: 74.6 years) and 6,938 controls[58,59]. A moderate but significant burden of (coding and non-coding) deletions was associated with AD status overall and appeared higher for rarest events (singletons). Some CNVs affected known AD genes including one coding partial deletion of *SORL1* and 5 coding partial deletions of *ABCA7* (all in cases). Interestingly, some non-coding structural variants, which are not detectable by exome sequencing, were also prioritized, suggesting that increasing sample sizes with WGS data should unveil novel associations. Novel long read sequencing technologies may also offer new opportunities, enabling the analysis of CNVs along with mobile element insertions, repeat variations, and balanced structural variants with an unprecedented accuracy[60,61], but sequencing costs remain high, limiting the sample sizes. In addition, we did not study the X chromosome, due to the complexity of the analysis models. Specific analysis of this chromosome may reveal potential variants associated with AD[62].

### ADDITIONAL ACKNOWLEDGEMENTS

*SURF supercomputer facility*

Part of the work in this manuscript was carried out on the Cartesius supercomputer, which is embedded in the Dutch national e-infrastructure with the support of SURF Cooperative. Computing hours were granted in 2016, 2017, 2018 and 2019 to H. Holstege by the Dutch Research Council (project name: '100plus'; project numbers 15318 and 17232).

*ADES-FR*

This study was funded by grants from the Clinical Research Hospital Program from the French ministry of Health (GMAJ, PHRC, 2008/067), the CNR-MAJ, the JPND PERADES, Equipe FRM DEQ20170336711, and Fondation Alzheimer (ECASCAD study). This research was supported by the Laboratory of Excellence GENMED (Medical Genomics) grant no. ANR-10-LABX-0013 managed by the National Research Agency (ANR) part of the Investment for the Future program. This work was also supported by Foundation Alzheimer, the Institut Pasteur de Lille, Inserm, the Haut-de-France and Lille Métropole Communauté Urbaine council, and the French government's LABEX (laboratory of excellence program investment for the future) DISTALZ grant (Development of Innovative Strategies for a Transdisciplinary approach to Alzheimer's disease). The 3C Study supports are listed on the Study Website (www.three-city-study.com). This work is a collaboration between CEADRF-Jacob-CNRGH-CHU de Rouen. This work did benefit from the support of the France Génomique National infrastructure, funded as part of the "Investissements d'Avenir" program managed by the Agence Nationale pour la Recherche (contract ANR-10-INBS-09).

*AgeCoDe-UKBonn*

The AgeCoDe cohort was funded in part by the German Federal Ministry of Education and Research (BMBF) (grants KNDD 01GI0710, 01GI0711, 01GI0712, 01GI0713, 01GI0714, 01GI0715, 01GI0716, 01ET1006B). Sequencing of AgeCoDe sample was in part funded by the German Research Foundation (DFG) grant RA 1971/6-1 to Alfredo Ramirez.

Barcelona- SPIN

Support for Jordi Clarimon provided by Maratón RTVE (Spain). Support for Oriol Dols provided by the Association for Frontotemporal Degeneration (Clinical Research Postdoctoral Fellowship, AFTD).

AC-EMC

Exome sequencing was funded by Alzheimer Nederland.

ERF

The ERF study as a part of EUROSPAN (European Special Populations Research Network) was supported by European Commission FP6 STRP grant number 018947 (LSHG-CT-2006-01947) and also received funding from the European Community's Seventh Framework Programme (FP7/2007-2013)/grant agreement HEALTH-F4- 2007-201413 by the European Commission under the programme "Quality of Life and Management of the Living Resources" of 5th Framework Programme (no. QLG2-CT-2002- 01254). High-throughput analysis of the ERF data was supported by a joint grant from the Netherlands Organization for Scientific Research and the Russian Foundation for Basic Research (NWO-RFBR 047.017.043).

Rotterdam Study

The generation and management of the exome sequencing data for the Rotterdam Study was executed by the Human Genotyping Facility of the Genetic Laboratory of the Department of Internal Medicine, Erasmus MC, the Netherlands. The Rotterdam Study is funded by Erasmus Medical Center and Erasmus University, Rotterdam, the Netherlands Organization for Health Research and Development (ZonMw), the Research Institute for diseases in the Elderly (RIDE) (014-93-015; RIDE2), the Ministry of Education, Culture and Science, the Ministry for Health, Welfare and Sports, the European Commission (DG XII), and the municipality of Rotterdam. Genetic data sets are also supported by the Netherlands Organization of Scientific Research NWO Investments (175.010.2005.011, 911-03-012), the Genetic Laboratory of the Department of Internal Medicine, Erasmus MC, and the Netherlands Genomics Initiative (NGI)/Netherlands Organization for Scientific Research (NWO), the Netherlands Consortium for Healthy Aging (NCHA), project 050-060-810, and by a Complementation Project of the Biobanking and Biomolecular Research Infrastructure Netherlands (BBMRI-NL; www.bbmri.nl ; project number CP2010-41). We thank Mr. Pascal Arp, Ms. Mila Jhamai, Mr. and Marijn Verkerk, for their help in creating the RS-Exome Sequencing database.

ADC-Amsterdam

We thank all study participants and all personnel involved in data collection for the contributing studies. Research of Alzheimer center Amsterdam is part of the neurodegeneration research program of Amsterdam Neuroscience. Alzheimer Center Amsterdam is supported by Stichting Alzheimer Nederland and Stichting VUmc fonds. The clinical database structure was developed with funding from Stichting Dioraphte. This work was supported by Stichting Alzheimer Nederland (WE.09-2014-06, WE.05-2010-06); Stichting Dioraphte; Internationale Stichting Alzheimer Onderzoek (#11519); JPNDPERADES (ZonMw 733051022): Centralized Facility for Sequence to Phenotype analyses (ZonMW 9111025); Netherlands Consortium for Healthy Aging (NCHA 050-060-810); Biobanking and Biomolecular Research Infrastructure Netherlands (BBMRI-NL CP2010-41); Netherlands Genomics Initiative (NGI)/NWO. This study is further supported by ABOARD, a public-private partnership receiving funding from ZonMW (#73305095007) and Health~Holland, Topsector Life Sciences & Health (PPP-allowance; #LSHM20106). This research is performed by using data from the Parelsnoer Institute an initiative of the Dutch Federation of University Medical Centres (www.parelsnoer.org).

100-plus Study

Cohort collection and exome sequencing of the 100-plus Study cohort was supported by Stichting Alzheimer Nederland (WE.09-2014-03); HorstingStuit Foundation, VUmc Foundation, and the

Dioraphte Foundation (Project 17020403), Memorabel (ZonMW project number #733050814, #733050512) and Stichting VUmcFonds. Additional support is from ABOARD, a public-private partnership receiving funding from ZonMW (#73305095007) and Health~Holland, Topsector Life Sciences & Health (PPP-allowance; #LSHM20106).

EMIF-AD 90+

The EMIF-AD 90+ Study was funded by the EU/EFPIA Innovative Medicines Initiative Joint Undertaking EMIF grant agreement no. 115372.

CBC: Control Brain Consortium

This work was supported by the UK Dementia Research Institute which receives its funding from DRI Ltd, funded by the UK Medical Research Council, Alzheimer's Society and Alzheimer's Research UK, Medical Research Council (award number MR/N026004/1). Wellcome Trust Hardy (award number 202903/Z/16/Z), Dolby Family Fund; National Institute for Health Research University College London Hospitals Biomedical Research Centre; BRCNIHR Biomedical Research Centre at University College London Hospitals NHS Foundation Trust and University College London. J. Hardy was supported by the Dolby Foundation and the JPND PERADES. J.B. and R.G were supported by the National Institute on Aging of the National Institutes of Health under Award Number R01AG067426. The content is solely the responsibility of the authors and does not necessarily represent the official views of the National Institutes of Health.

PERADES

We thank all individuals who participated in the study. We also want to express our gratitude to the MRC Centre Core Team for the laboratory support and the Advanced Research Computing at Cardiff University (ARCCA) for the computational support. Cardiff University was supported by the Medical Research Council. Cardiff University was also supported by the European Joint Programme for Neurodegenerative Disease, Alzheimer's Research UK, the Welsh Assembly Government, and a donation from the Moondance Charitable Foundation. Cardiff University acknowledges the support of the UK Dementia Research Institute, which receives its funding from UK DRI Ltd, funded by the UK Medical Research Council, Alzheimer's Society and Alzheimer's Research UK. Cambridge University acknowledges support from the MRC. The University of Southampton acknowledges support from the Alzheimer's Society. ARUK provided support to Nottingham University. Join Dementia Research (JDR) is funded by the Department of Health and delivered by the National Institute for Health Research in partnership with Alzheimer Scotland, Alzheimer's Research UK and Alzheimer's Society. IRCCS Santa Lucia Foundation acknowledges the Italian Ministry of Health for financial support (IMH_RC) of this study. The Centro de Biologia de Molecular Severo Ochoa (CSIS-UAM), CIBERNED, Instituto de Investigacion Sanitaria la Paz, University Hospital La Paz and the Universidad Autonoma de Madrid were supported by grants from the Ministerio de Educacion y Ciencia and the Ministerio de Sanidad y Consumo (Instituto de Salud Carlos III), and an institutional grant of the Fundacion Ramon Areces to the CMBSO. Thanks to I. Sastre and Dr A Martinez-Garcia for DNA preparation, and Drs P Gil and P Coria for their recruitment efforts. Department of Neurology, University Hospital Mutua de Terrassa, Terrassa, Barcelona, Spain was supported by CIBERNED, Centro de Investigacion Biomedica en Red de Enfermedades Neurodegenerativas, Instituto de Salud Carlos III, Madrid Spain and acknowledges Maria A Pastor (Department of Neurology, University of Navarra Medical School and Neuroimaging

Laboratory, Center for Applied Medical Research, Pamplona, Spain), Manuel Seijo-Martinez (Department of Neurology, Hospital do Salnes, Pontevedra, Spain), Ramon Rene, Jordi Gascon and Jaume Campdelacreu (Department of Neurology, Hospital de Bellvitage, Barcelona, Spain) for providing DNA samples. Hospital de la Sant Pau, Universitat Autonoma de Spain acknowledges support from the Spanish Ministry of Economy and Competitiveness (grant number PI12/01311), and from Generalitat de Catalunya (2014SGR-235). The Santa Lucia Foundation and the Fondazione Ca' Granda IRCCS Ospedale Policlinico, Italy, acknowledge the Italian Ministry of Health (grant RC 10.11.12.13/A)

UCL-DRC EOAD

This work was supported by the Medical Research Council (UK), the Biomedical Research Centre at University College London Hospitals NHS Foundation Trust and charitable donations to the UCL Dementia Research Centre.

ADSP

The Alzheimer's Disease Sequencing Project (ADSP) is comprised of two Alzheimer's Disease (AD) genetics consortia and three National Human Genome Research Institute (NHGRI) funded Large Scale Sequencing and Analysis Centers (LSAC). The two AD genetics consortia are the Alzheimer's Disease Genetics Consortium (ADGC) funded by NIA (U01 AG032984), and the Cohorts for Heart and Aging Research in Genomic Epidemiology (CHARGE) funded by NIA (R01 AG033193), the National Heart, Lung, and Blood Institute (NHLBI), other National Institute of Health (NIH) institutes and other foreign 104 governmental and non-governmental organizations. The Discovery Phase analysis of sequence data is supported through UF1AG047133 (to Drs. Schellenberg, Farrer, Pericak- Vance, Mayeux, and Haines); U01AG049505 to Dr. Seshadri; U01AG049506 to Dr. Boerwinkle; U01AG049507 to Dr. Wijsman; and U01AG049508 to Dr. Goate and the Discovery Extension Phase analysis is supported through U01AG052411 to Dr. Goate, U01AG052410 to Dr. Pericak-Vance and U01 AG052409 to Drs. Seshadri and Fornage, U54 AG052427 to Drs. Schellenberg and Wang, and R01 AG054060 to Dr Naj. The ADGC cohorts include: Adult Changes in Thought (ACT) (UO1 AG006781, UO1 HG004610, UO1 HG006375, U01 HG008657), the Alzheimer's Disease Centers (ADC) ( P30 AG019610, P30 AG013846, P50 AG008702, P50 AG025688, P50 AG047266, P30 AG010133, P50 AG005146, P50 AG005134, P50 AG016574, P50 AG005138, P30 AG008051, P30 AG013854, P30 AG008017, P30 AG010161, P50 AG047366, P30 AG010129, P50 AG016573, P50 AG016570, P50 AG005131, P50 AG023501, P30 AG035982, P30 AG028383, P30 AG010124, P50 AG005133, P50 AG005142, P30 AG012300, P50 AG005136, P50 AG033514, P50 AG005681, and P50 AG047270), the Chicago Health and Aging Project (CHAP) (R01 AG11101, RC4 AG039085, K23 AG030944), Indianapolis Ibadan (R01 AG009956, P30 AG010133), the Memory and Aging Project (MAP) ( R01 AG17917), Mayo Clinic (MAYO) (R01 AG032990, U01 AG046139, R01 NS080820, RF1 AG051504, P50 AG016574), Mayo Parkinson's Disease controls (NS039764, NS071674, 5RC2HG005605), University of Miami (R01 AG027944, R01 AG028786, R01 AG019085, IIRG09133827, A2011048), the Multi-Institutional Research in Alzheimer's Genetic Epidemiology Study (MIRAGE) (R01 AG09029, R01 AG025259), the National Cell Repository for Alzheimer's Disease (NCRAD) (U24 AG21886), the National Institute on Aging Late Onset Alzheimer's Disease Family Study (NIA- LOAD) (R01 AG041797), the Religious Orders Study (ROS) (P30 AG10161, R01 AG15819), the Texas Alzheimer's Research and Care Consortium (TARCC) (funded by the Darrell K Royal Texas Alzheimer's Initiative), Vanderbilt University/Case Western Reserve University (VAN/CWRU) (R01 AG019757, R01 AG021547, R01 AG027944, R01 AG028786, P01 NS026630, and

Alzheimer's Association), the Washington Heights-Inwood Columbia Aging Project (WHICAP) (RF1 AG054023), the University of Washington Families (VA Research Merit Grant, NIA: P50AG005136, R01AG041797, NINDS: R01NS069719), the Columbia University HispanicEstudio Familiar de Influencia Genetica de Alzheimer (EFIGA) (RF1 AG015473), the University of Toronto (UT) (funded by Wellcome Trust, Medical Research Council, Canadian Institutes of Health Research), and Genetic Differences (GD) (R01 AG007584). The CHARGE cohorts are supported in part by National Heart, Lung, and Blood Institute (NHLBI) infrastructure grant HL105756 (Psaty), RC2HL102419 (Boerwinkle) and the neurology working group is supported by the National Institute on Aging (NIA) R01 grant AG033193. This work was also supported by National Institute on Aging grants R01 AG048927 to Dr. Farrer, RF1 AG054080 to Dr. Beecham, U24 AG056270 to Dr. Mayeux, RF1 AG057519 to Dr. Farrer, U01 AG062602 to Dr. Farrer, R01 AG067501 to Dr. Mayeux, and U19 AG068753 to Dr. Farrer. The CHARGE cohorts participating in the ADSP include the following: Austrian Stroke Prevention Study (ASPS), ASPS-Family study, and the Prospective Dementia Registry- Austria (ASPS/PRODEM-Aus), the Atherosclerosis Risk in Communities (ARIC) Study, the Cardiovascular Health Study (CHS), the Erasmus Rucphen Family Study (ERF), the Framingham Heart Study (FHS), and the Rotterdam Study (RS). ASPS is funded by the Austrian Science Fond (FWF) grant number P20545-P05 and P13180 and the Medical University of Graz. The ASPS-Fam is funded by the Austrian Science Fund (FWF) project I904),the EU Joint Programme - Neurodegenerative Disease Research (JPND) in frame of the BRIDGET project (Austria, Ministry of Science) and the Medical University of Graz and the Steiermärkische Krankenanstalten Gesellschaft. PRODEM-Austria is supported by the Austrian Research Promotion agency (FFG) (Project No. 827462) and by the Austrian National Bank (Anniversary Fund, project 15435. ARIC research is carried out as a collaborative study supported by NHLBI contracts (HHSN268201100005C, HHSN268201100006C, HHSN268201100007C, HHSN268201100008C, HHSN268201100009C, HHSN268201100010C, HHSN268201100011C, and HHSN268201100012C). Neurocognitive data in ARIC is collected by U01 2U01HL096812, 2U01HL096814, 2U01HL096899, 2U01HL096902, 2U01HL096917 from the NIH (NHLBI, NINDS, NIA and NIDCD), and with previous brain MRI examinations funded by R01- HL70825 from the NHLBI. CHS research was supported by contracts HHSN268201200036C, HHSN268200800007C, N01HC55222, N01HC85079, N01HC85080, N01HC85081, N01HC85082, N01HC85083, N01HC85086, and grants U01HL080295 and U01HL130114 from the NHLBI with additional contribution from the National Institute of Neurological Disorders and Stroke (NINDS). Additional support was provided by R01AG023629, R01AG15928, and R01AG20098 from the NIA. FHS research is supported by NHLBI contracts N01-HC-25195 and HHSN268201500001I. This study was also supported by additional grants from the NIA (R01s AG054076, AG049607 and AG033040 and NINDS (R01 NS017950). The ERF study as a part of EUROSPAN (European Special Populations Research Network) was supported by European Commission FP6 STRP grant number 018947 (LSHG-CT-2006-01947) and also received funding from the European Community's Seventh Framework Programme (FP7/2007- 2013)/grant agreement HEALTH-F4- 2007-201413 by the European Commission under the programme "Quality of Life and Management of the Living Resources" of 5th Framework Programme (no. QLG2-CT-2002- 01254). High-throughput analysis of the ERF data was supported by a joint grant from the Netherlands Organization for Scientific Research and the Russian Foundation for Basic Research (NWO-RFBR 047.017.043). The Rotterdam Study is funded by Erasmus Medical Center and Erasmus University, Rotterdam, the Netherlands Organization for Health Research and Development (ZonMw), the Research Institute for Diseases in the Elderly (RIDE), the Ministry of Education, Culture and Science, the Ministry for Health, Welfare and Sports, the European Commission (DG XII), and the municipality of Rotterdam. Genetic data sets are also supported by the Netherlands Organization of Scientific Research NWO Investments (175.010.2005.011, 911-03-012), the Genetic Laboratory of the Department of Internal Medicine, Erasmus MC, the Research Institute for Diseases in the Elderly (014-93-015; RIDE2), and the

Netherlands Genomics Initiative (NGI)/Netherlands Organization for Scientific Research (NWO) Netherlands Consortium for Healthy Aging (NCHA), project 050-060-810. All studies are grateful to their participants, faculty and staff. The content of these manuscripts is solely the responsibility of the authors and does not necessarily represent the official views of the National Institutes of Health or the U.S. Department of Health and Human Services. The four LSACs are: the Human Genome Sequencing Center at the Baylor College of Medicine (U54 HG003273), the Broad Institute Genome Center (U54HG003067), The American Genome Center at the Uniformed Services University of the Health Sciences (U01AG057659), and the Washington University Genome Institute (U54HG003079). Biological samples and associated phenotypic data used in primary data analyses were stored at Study Investigators institutions, and at the National Cell Repository for Alzheimer's Disease (NCRAD, U24AG021886) at Indiana University funded by NIA. Associated Phenotypic Data used in primary and secondary data analyses were provided by Study Investigators, the NIA funded Alzheimer's Disease Centers (ADCs), and the National Alzheimer's Coordinating Center (NACC, U01AG016976) and the National Institute on Aging Genetics of Alzheimer's Disease Data Storage Site (NIAGADS, U24AG041689) at the University of Pennsylvania, funded by NIA This research was supported in part by the Intramural Research Program of the National Institutes of health, National Library of Medicine. Contributors to the Genetic Analysis Data included Study Investigators on projects that were individually funded by NIA, and other NIH institutes, and by private U.S. organizations, or foreign governmental or nongovernmental organizations. Data used in the preparation of this article were obtained from the Alzheimer's Disease Neuroimaging Initiative (ADNI) database (adni.loni.usc.edu). The ADNI was launched in 2003 as a public-private partnership, led by Principal Investigator Michael W. Weiner, MD. The primary goal of ADNI has been to test whether serial magnetic resonance imaging (MRI), positron emission tomography (PET), other biological markers, and clinical and neuropsychological assessment can be combined to measure the progression of mild cognitive impairment (MCI) and early Alzheimer's disease (AD). For up-to-date information, see www.adni-info.org. Data collection and sharing for this project was funded by the Alzheimer's Disease Neuroimaging Initiative (ADNI) (National Institutes of Health Grant U01 AG024904) and DOD ADNI (Department of Defense award number W81XWH-12-2-0012). ADNI is funded by the National Institute on Aging, the National Institute of Biomedical Imaging and Bioengineering, and through generous contributions from the following: AbbVie, Alzheimer's Association; Alzheimer's Drug Discovery Foundation; Araclon Biotech; BioClinica, Inc.; Biogen; Bristol-Myers Squibb Company; CereSpir, Inc.; Cogstate; Eisai Inc.; Elan Pharmaceuticals, Inc.; Eli Lilly and Company; EuroImmun; F. Hoffmann-La Roche Ltd and its affiliated company Genentech, Inc.; Fujirebio; GE Healthcare; IXICO Ltd.; Janssen Alzheimer Immunotherapy Research & Development, LLC.; Johnson & Johnson Pharmaceutical Research & Development LLC.; Lumosity; Lundbeck; Merck & Co., Inc.; Meso Scale Diagnostics, LLC.; NeuroRx Research; Neurotrack Technologies; Novartis Pharmaceuticals Corporation; Pfizer Inc.; Piramal Imaging; Servier; Takeda Pharmaceutical Company; and Transition Therapeutics. The Canadian Institutes of Health Research is providing funds to support ADNI clinical sites in Canada. Private sector contributions are facilitated by the Foundation for the National Institutes of Health (www.fnih.org). The grantee organization is the Northern California Institute for Research and Education, and the study is coordinated by the Alzheimer's Therapeutic Research Institute at the University of Southern California. ADNI data are disseminated by the Laboratory for Neuro Imaging at the University of Southern California.

### SUPPLEMENTARY FIGURES

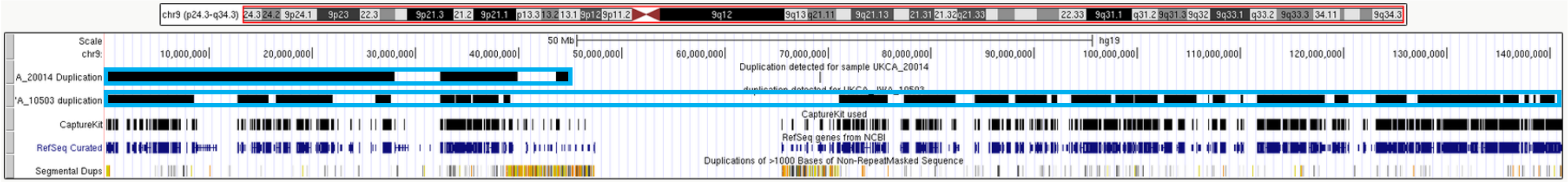

**Figure S1. Example of a large CNV on chromosome 9 suggesting a blood-specific event (probable clonal hematopoiesis)**

This figure was obtained from the UCSC genome browser and represents large mosaic duplications detected in two samples from the PERADES dataset on chromosome 9 (GRCh37/hg19). The first two rows display duplications detected across chromosome 9. Black rectangles indicate duplications as defined by CANOES. Blue rectangles indicate how multiple calls should be merged as one unique event. The third line indicates the position of capture kit targets (both samples have the same capture kit). The fourth line indicates genes position according to RefSeq. Finally, the last line indicates positions of segmental duplication regions.

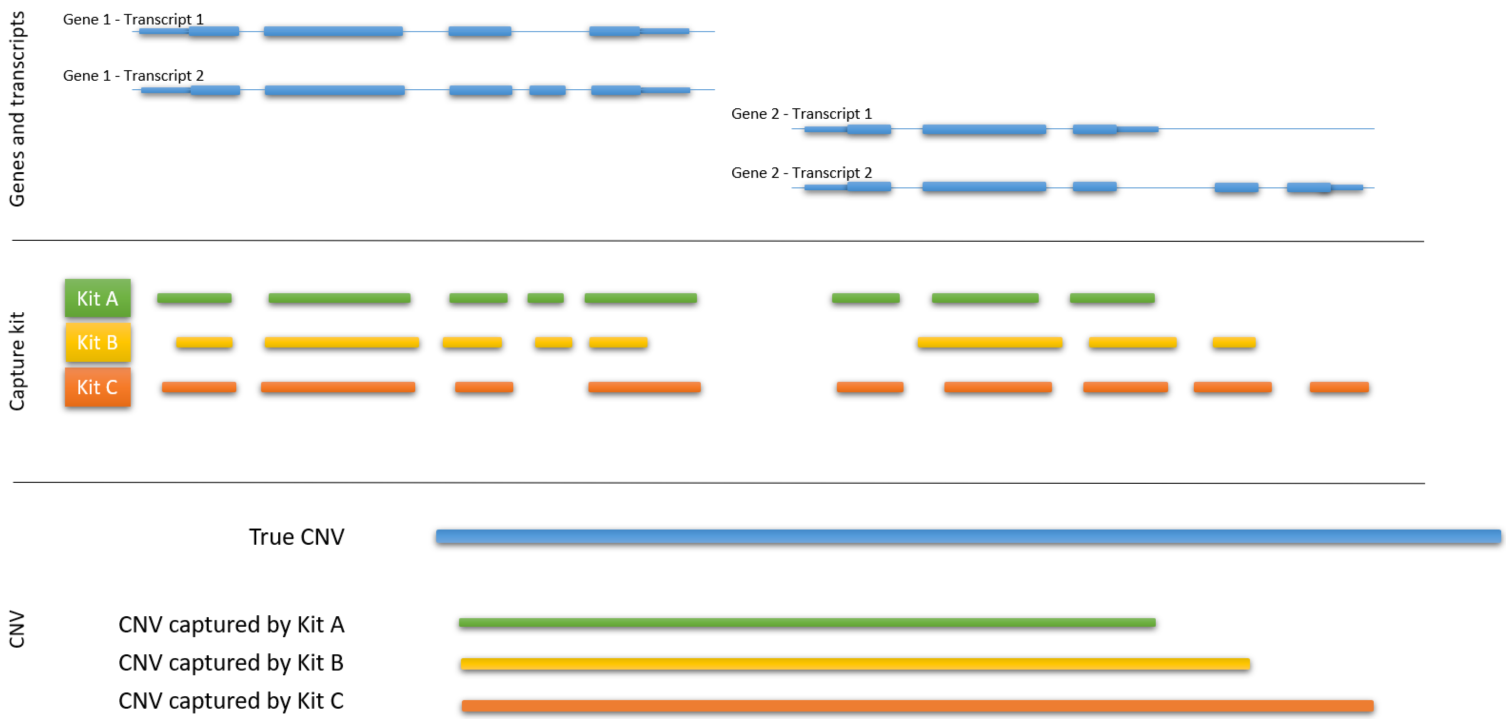

**Figure S2. Theoretical example of definition of CNV coordinates according to transcript and capture kit coordinates.**

On the top, a theoretical example of two genes with two transcripts each. Larger blue rectangles represent exons whereas thinner rectangles represent non coding regions.
On the middle, an example of a difference of captured zone between different kits.
On the bottom, the theoretical true CNV, in blue, and the coordinates attributed by each capture kit to this CNV.

In this example, using only transcript coordinates leads to a CNV considered as encompassing fully Gene 2 – transcripts 1 and 2 only for data captured by kit C; whereas, using our own transcript coordinates (defined by available targets encompassing the transcript), this CNV is considered as fully affecting Gene 2 – transcripts 1 and 2 whatever the capture kit used. For each capture kit, Gene 1 is supposed to be affected partially.

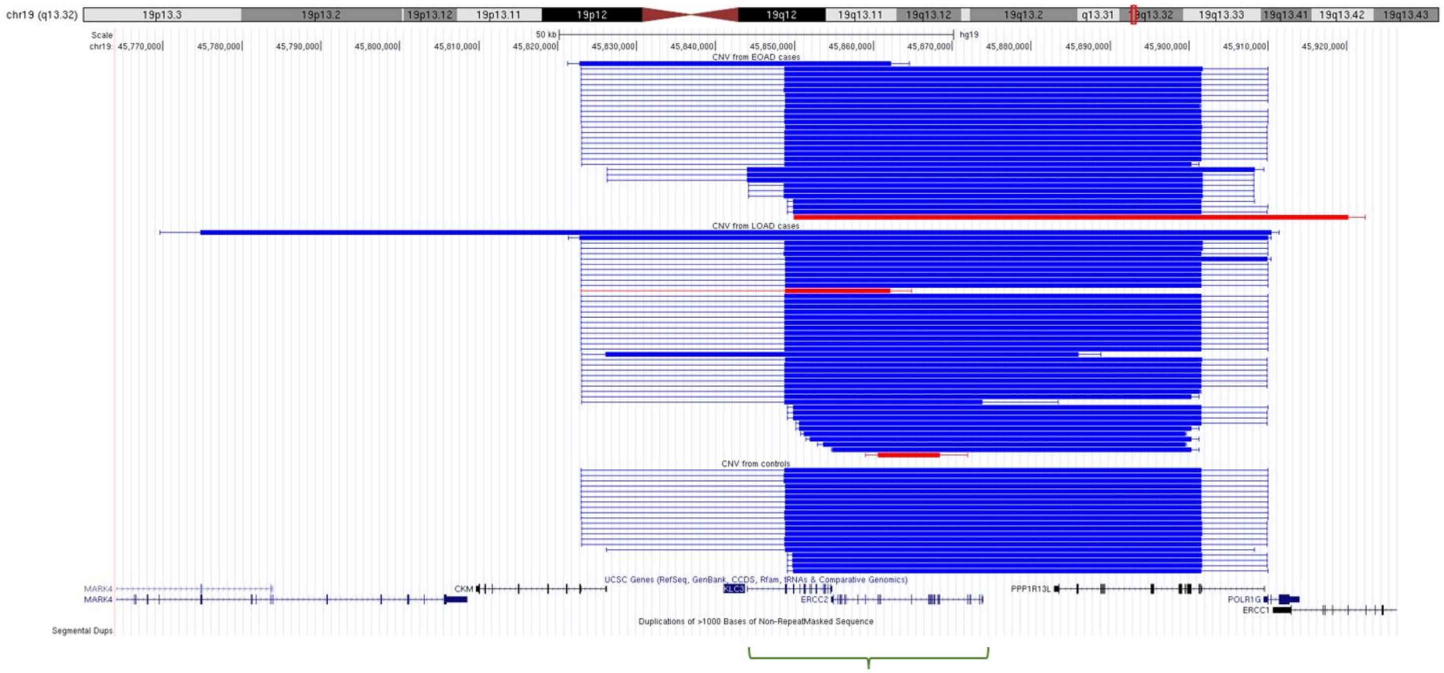

**Figure S3.** ***ERCC2-KLC3*** **locus on chr19 with CNVs displayed on the UCSC genome browser**

In red: deletions, In blue: duplications

Large areas represent coordinates of CNVs as detected by CANOES. Thinner areas show breakpoints uncertainties, i.e. regions between two targets of the sample-specific capture kit. Genes are indicated below, and the regions of segmental duplications appear at the bottom part (none here). The green area indicates the locus identified in our study.

Note that most of the duplications share similar coordinates, also encompassing the *PPP1R13L* gene but transcripts of this gene were not considered in the analysis because they were not predicted to be entirely duplicated. This locus is on chromosome 19q13.32, ~440-kb away from the APOE gene and duplications were significantly more frequently carried by APOE4+ individuals, independently of the disease status (Table S12), Consistently, the association signal did not remain significant after adjusting for APOE-ε4 dosage (OR=1.58 [0.85; 2.99], p=0.148) while other loci remained unaffected (Table S10).

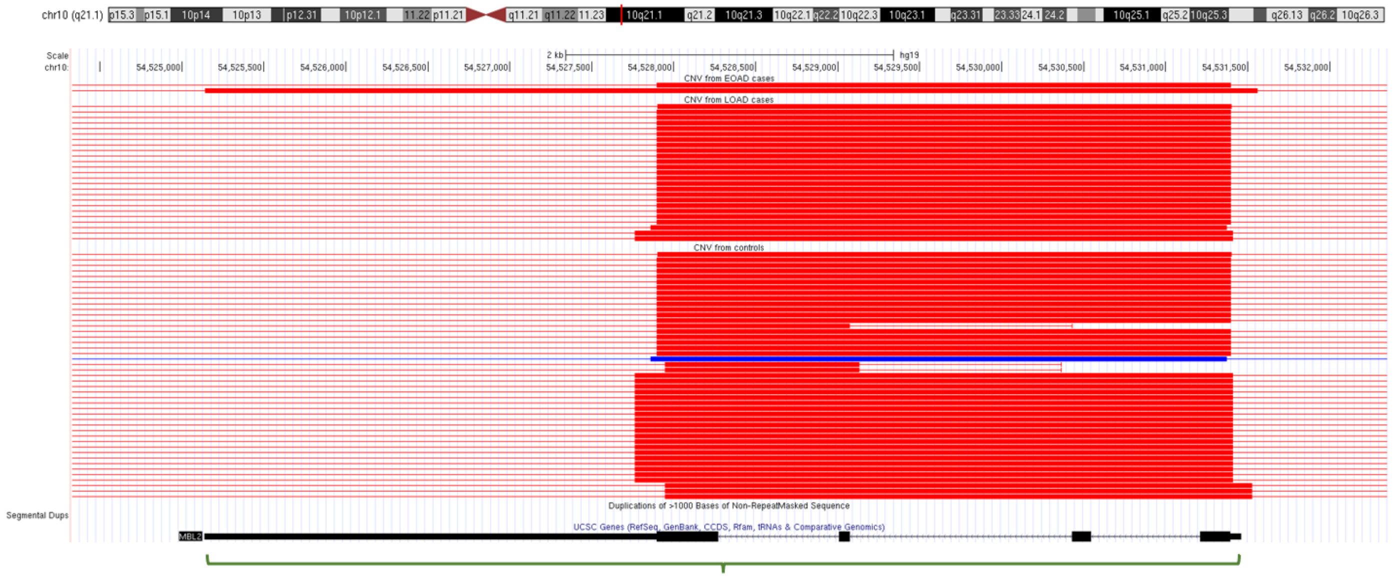

**Figure S4. *MBL2* locus with CNVs displayed on the UCSC genome browser**

In red: deletions
In blue: duplications

Large areas represent coordinates of CNVs as detected by CANOES. Thinner areas show breakpoints uncertainties, i.e. regions between two targets of the sample-specific capture kit.
Genes are indicated below, and the regions of segmental duplications appear at the bottom part (none here).

The green area indicates the locus identified in our study

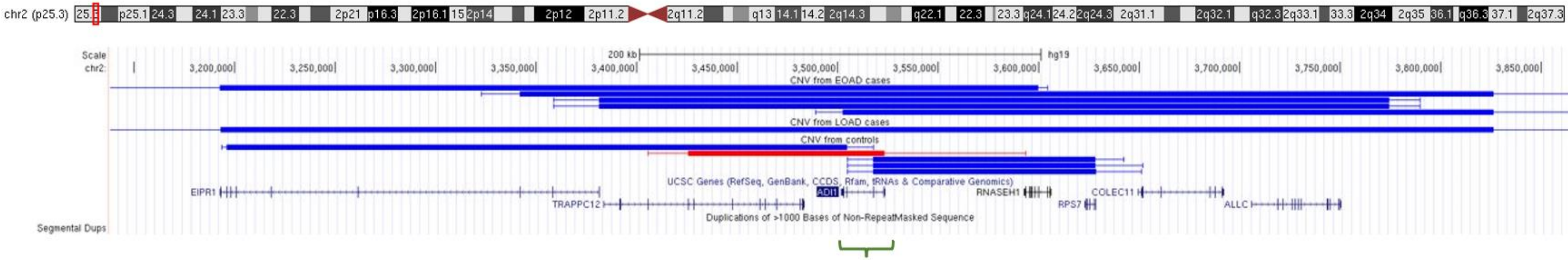

**Figure S5.** ***ADI1*** **locus with CNVs displayed on the UCSC genome browser**

In red: deletions
In blue: duplications

Large areas represent coordinates of CNVs as detected by CANOES. Thinner areas show breakpoints uncertainties, i.e. regions between two targets of the sample-specific capture kit.
Genes are indicated below, and the regions of segmental duplications appear at the bottom part (none here).
The green area indicates the locus identified in our study.

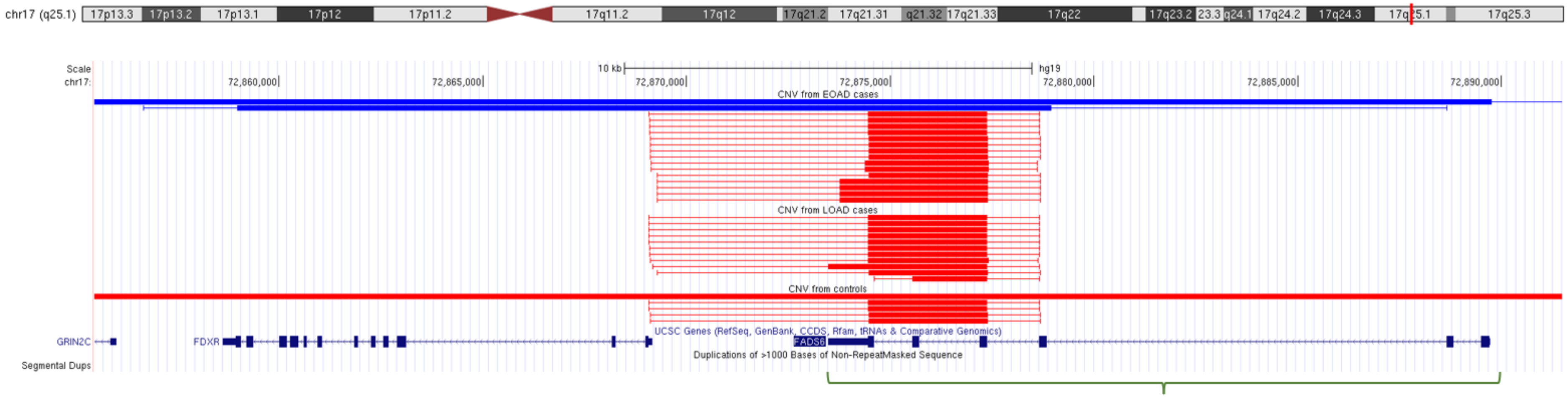

**Figure S6. *FADS6* locus with CNVs displayed on the UCSC genome browser**

In red: deletions
In blue: duplications

Large areas represent coordinates of CNVs as detected by CANOES. Thinner areas show breakpoints uncertainties, i.e. regions between two targets of the sample-specific capture kit.
Genes are indicated below, and the regions of segmental duplications appear at the bottom part (none here).

The green area indicates the locus identified in our study.

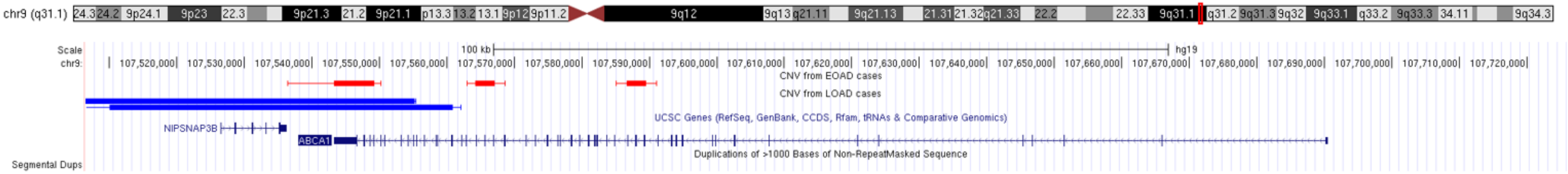

**Figure S7. *ABCA1* locus with CNVs displayed on the UCSC genome browser**

In red: deletions
In blue: duplications

Large areas represent coordinates of CNVs as detected by CANOES. Thinner areas show breakpoints uncertainties, i.e. regions between two targets of the sample-specific capture kit.
Genes are indicated below, and the regions of segmental duplications appear at the bottom part (none here).

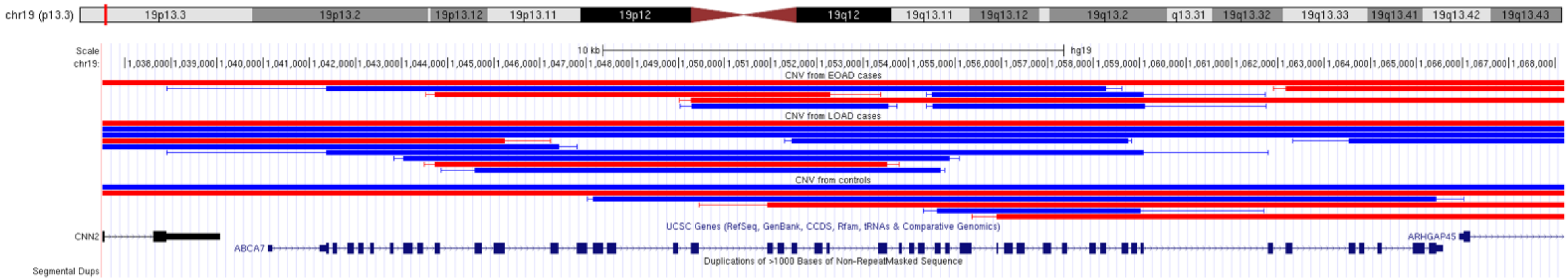

**Figure S8. *ABCA7* locus with CNVs displayed on the UCSC genome browser**

In red: deletions
In blue: duplications

Large areas represent coordinates of CNVs as detected by CANOES. Thinner areas show breakpoints uncertainties, i.e. regions between two targets of the sample-specific capture kit.
Genes are indicated below, and the regions of segmental duplications appear at the bottom part (none here).

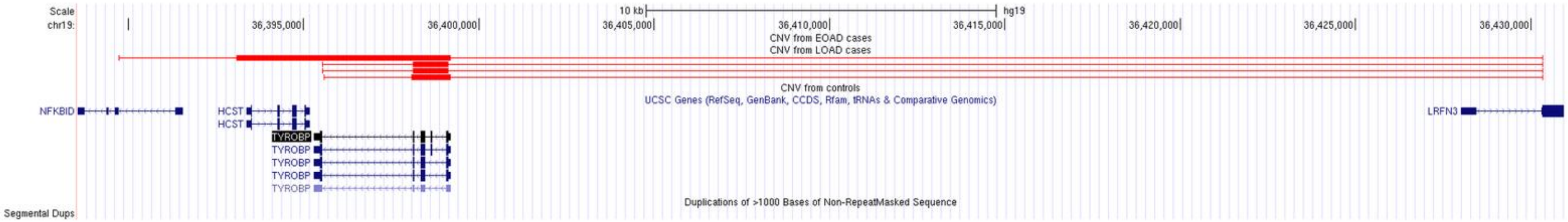

**Figure S9. *TYROBP* locus with CNVs displayed on the UCSC genome browser**

In red: deletions
In blue: duplications

Large areas represent coordinates of CNVs as detected by CANOES. Thinner areas show breakpoints uncertainties, i.e. regions between two targets of the sample-specific capture kit.
Genes are indicated below, and the regions of segmental duplications appear at the bottom part (none here).

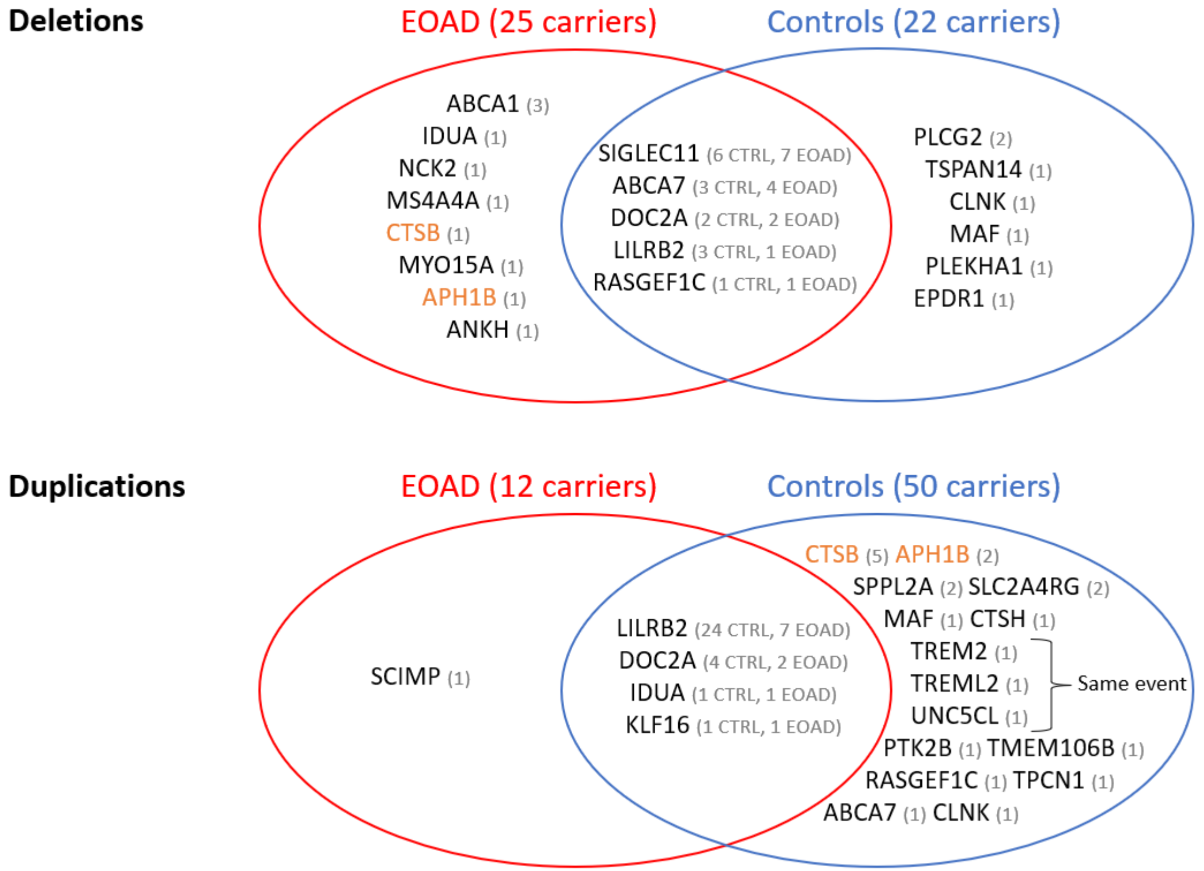

**Figure S10. CNVs affecting known AD-associated genes.**

Gray numbers corresponds to number of carriers of CNV encompassing the gene. In orange, genes with deletions in cases and duplications in controls, suggesting a dosage effect.

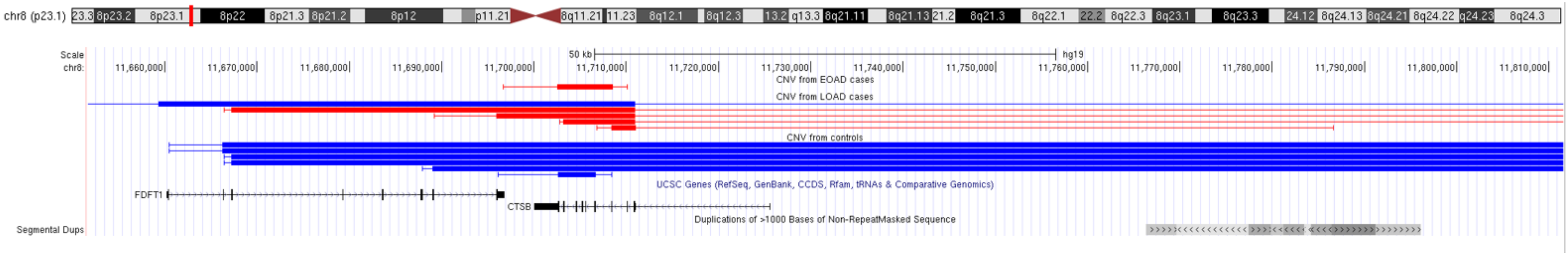

**Figure S11. *CTSB* locus with CNVs displayed on the UCSC genome browser**

In red: deletions
In blue: duplications

Large areas represent coordinates of CNVs as detected by CANOES. Thinner areas show breakpoints uncertainties, i.e. regions between two targets of the sample-specific capture kit.
Genes are indicated below, and the regions of segmental duplications appear at the bottom part.

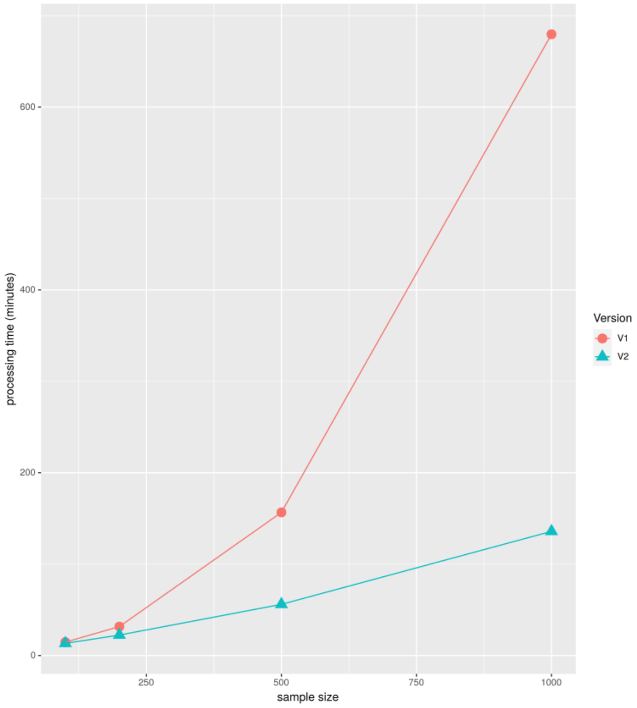

**Figure S12. Time required for the calling step of our pipeline with two versions of CANOES**

The two versions of CANOES differ only on the parallelization step, there is no differences on CNVs calling.

The same data have been processed using the same computational server with 32 threads and 192Gb of RAM. The parallelization used all threads, with the exception of the 1000 samples datasets, limited to 16 threads due to RAM limitation due to the matrix size**.**

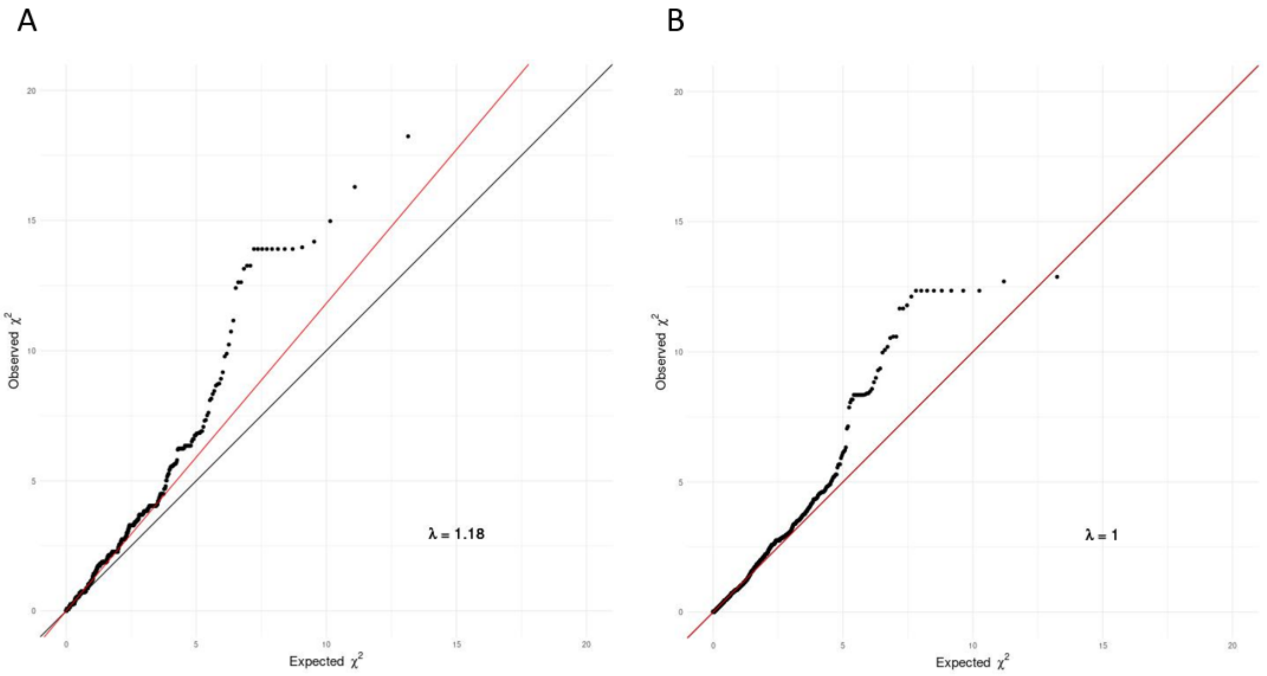

**Figure S13. QQ-plot for dosage analysis before and after adjustment for ancestry in EOAD versus Controls analysis.**

A. P-value qq-plot for main dosage analysis.
B. P-value qq-plot for ancestry adjusted dosage analysis.

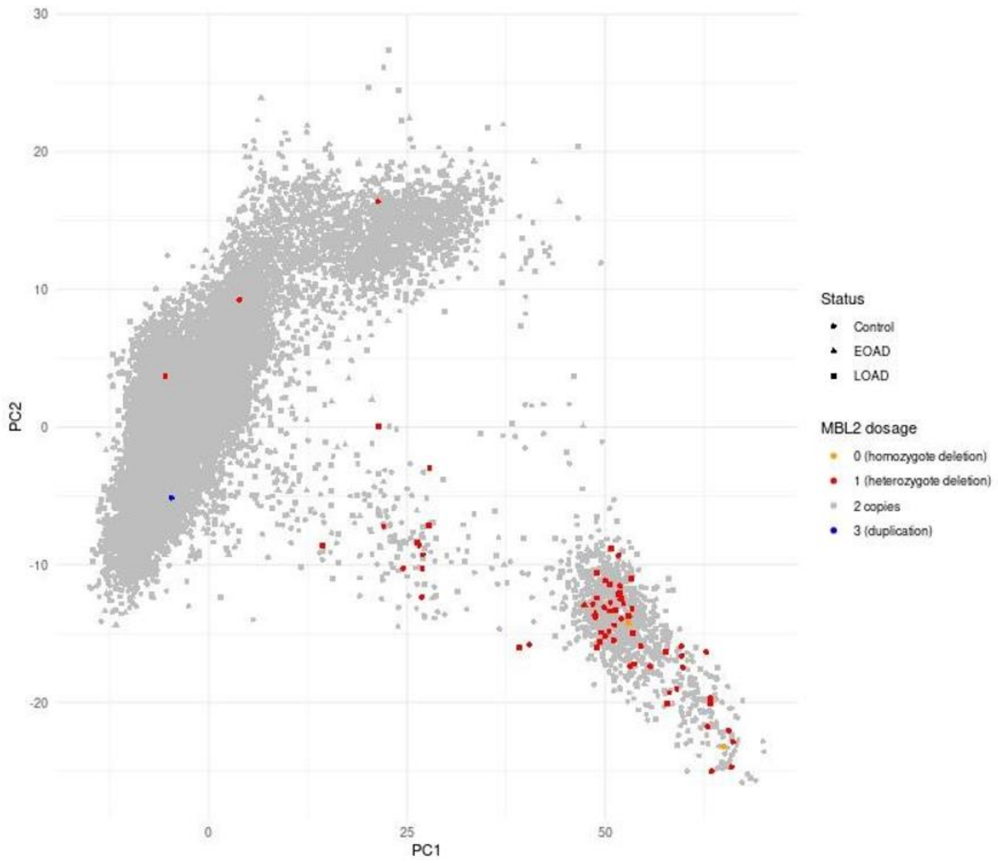

**Figure S14. Projection of *MBL2* CNV carriers on the first two ancestry components.**

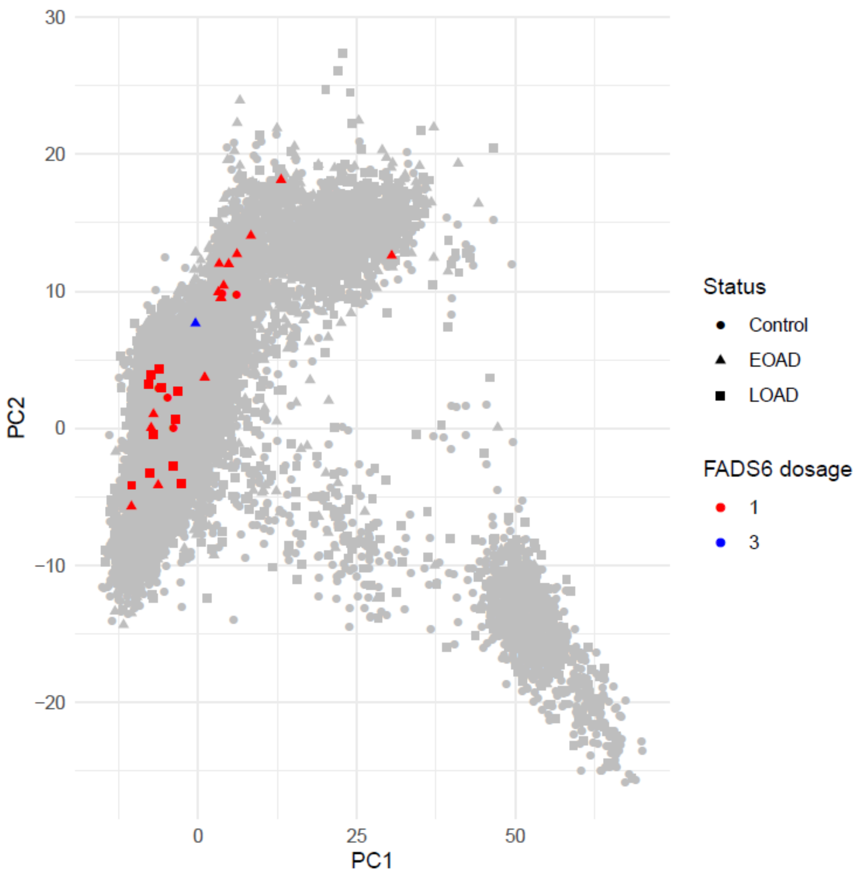

**Figure S15. Projection of *FADS6* CNV carriers on the first two ancestry components.**

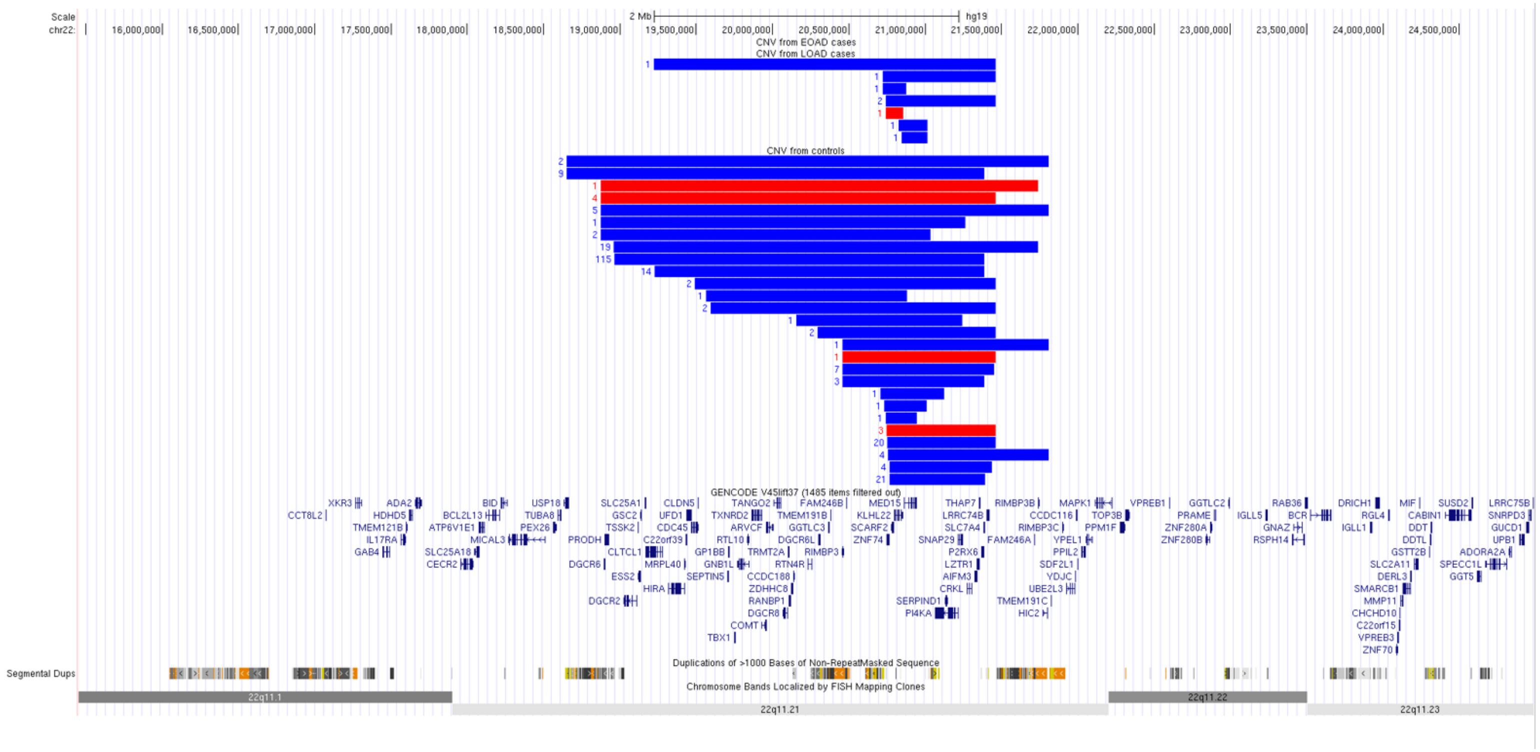

**Figure S16. 22q11.21 locus with CNVs from replication displayed on the UCSC genome browser**

In red: deletions
In blue: duplications

The number of occurrence of each CNV is indicated on the left
Genes are indicated below, and the regions of segmental duplications appear at the bottom part.

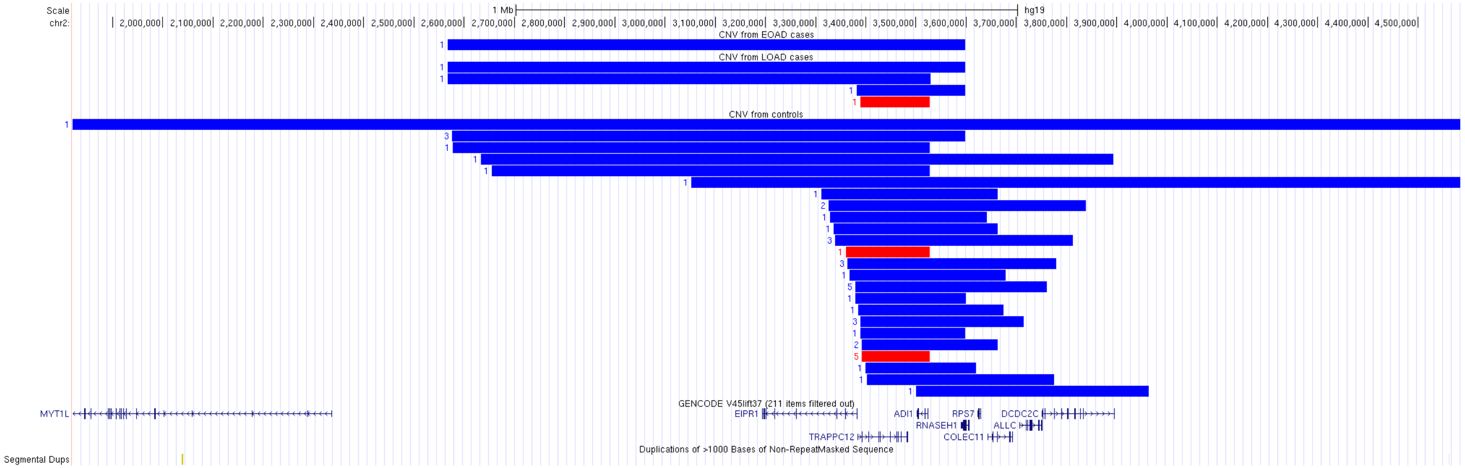

**Figure S17. *ADI1* locus with CNVs from replication displayed on the UCSC genome browser**

In red: deletions
In blue: duplications

The number of occurrence of each CNV is indicated on the left
Genes are indicated below, and the regions of segmental duplications appear at the bottom part.

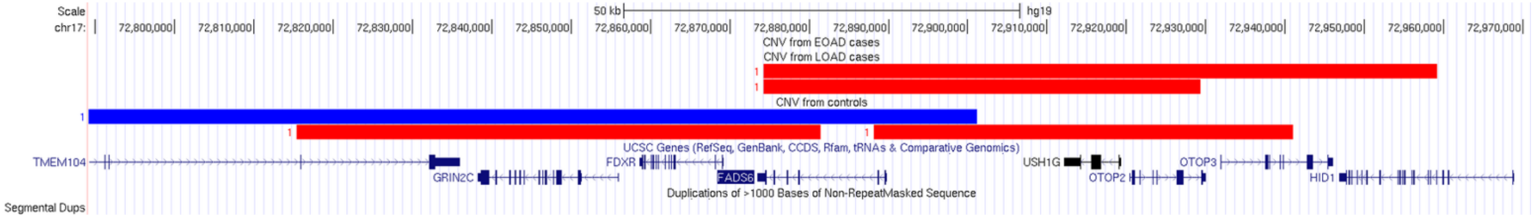

**Figure S18. *FADS6* locus with CNVs from replication displayed on the UCSC genome browser**

In red: deletions
In blue: duplications

The number of occurrence of each CNV is indicated on the left
Genes are indicated below, and the regions of segmental duplications appear at the bottom part.

### EXTENDED DATASET

**"Counts CNV per transcript and AD status.txt" is provided as a supplemental file.**

Counts of deletions and complete duplications per transcript after QC in the full dataset (summary statistics)

**"supplementary_Author_EADB.xlsx" is provided as a supplemental file.**

This file contains the complete list of EADB consortium members, as well as their respective affiliations.
